# Who are the Speech-Language Pathologists of the Future? Results of a national demographic survey of Canadian SLP Students

**DOI:** 10.1101/2024.09.17.24313750

**Authors:** Emily Wood, Mariya Kika, Monika Molnar

**Affiliations:** Department of Speech-Language Pathology, University of Toronto; Rehabilitation Sciences Institute, University of Toronto

## Abstract

Speech-language pathologists (SLPs) are experts in communication and swallowing who work with individuals across the lifespan. Prior demographic surveys in English-speaking countries (e.g., Canada and the United States) suggest that SLPs’ gender, racial, linguistic, and cultural and socioeconomic backgrounds may not be aligned with those of their clientele. The aim of this study is to describe the demographic characteristics of current SLP students, to gauge whether the demographic composition of future clinicians is changing and is aligned with the population they are trained to serve. We designed an anonymous online survey, in the Canadian context, that enabled us to compare current SLP student demographics with statistics available about the target population at large. This survey was disseminated to all SLP students enrolled in an accredited institution in 2024. Participants answered questions about their age, sex and gender, linguistic, racial and cultural identities, and socioeconomic status. More than half (N=525, 53%) of currently enrolled SLP students completed the survey. Results indicate that SLP students are overwhelmingly cisgender females. SLP students spoke 48 unique languages, and while most were bi or multilingual, few felt competent enough in languages other than English and French to engage in clinical service delivery. There were 151 unique racial and ethnic identities reported, with the largest proportion of students identifying as “North American.” Culturally most SLPs identified as “Canadian.” Very few students identified as Black or Indigenous racially or culturally. Most respondents reported a higher-than-average total household income. Findings suggest that the demographic composition of current SLP students is more diverse than those currently practicing, however there is still over-, and underrepresentation of certain populations. Implications for practice and suggestions for future research are discussed.

## Introduction

Communication, including spoken, written, and signed language, is a basic human right and is critical for participation in society – irrespective of linguistic and or cultural identity (United Nations General Assembly, 1949). Speech Language Pathologists (SLPs) serve clients ranging in age from newborns to the elderly and are the primary professionals responsible for providing assessment and intervention for communication and swallowing disorders (Speech-Language Pathology and Audiology Canada (SAC), 2016). Currently, graduate level SLP degrees can be obtained in several countries where the societal language is English (e.g., Australia, the United Kingdom, Canada and the United States) and linguistic and cultural diversity is common (e.g., Australian Bureau of Statistics, 2022; Office for National Statistics, 2022; Statistics Canada 2022d United States Census Bureau, 2022). In the current paper, we examine whether the future demographic composition of SLPs aligns with the demographic composition of the clientele they are trained to serve. To do so, we focused on the geographical area of Canada. Canada is an ideal test bed for this question, given that it provides SLP training to approximately 1000 students per year – a population size that is feasible to access, and it has recently published up-to-date detailed statistics about its residents’ demographic characteristics (Statistics Canada, 2022d).

There have been previous efforts to collect demographic information about practicing SLPs in other countries where English is the societal language and linguistic and cultural diversity is the norm. In the United States, the American Speech and Hearing Association (ASHA) has collected demographic information about its practicing members for several years. Of the over 200,000 SLPs who provided this information in 2023, 96.3% were females. Only 9.5% of members identified as either Asian, Black, Hawaiian / Pacific Islander, Mixed, American Indian / Alaska Native or Other in contrast to 38.4% of the American population at large who claim these racial identities (ASHA, 2023). ASHA has also partnered with the Council of Academic Programs in Communication Sciences and Disorders (CAPCSD) to collect demographic data of American SLP and audiology students since 2010. Results from the latest survey suggest that 27.4% of students have a racial identity other than Caucasian (CAPCSD, 2023). This number is much closer to the proportion of individuals with minority racial identities in the broader American population. This is promising, and potentially reflects efforts from American institutions to diversify the clinical workforce so that it better represents the population at large. However, ASHA’s member affiliate profile does not report data on languages status (e.g., mono/bilingual), languages spoken or language proficiency by SLPs or students. They report only whether SLPs identify as Hispanic or Latino (7.2%) or non-Hispanic or Latino (92.8%). No additional insights into the cultural identities of their membership are reported (ASHA, 2023).

In Australia, a national survey was disseminated to already practicing SLPs in 2021 (Nancarrow et al., 2023). Approximately 15% of those registered with their federal body responded. Like in the US, most respondents were females (99.6%). The most spoken languages were English, followed by Mandarin, Cantonese, Greek, Italian, Arabic and Spanish. The percentages of clinicians who spoke each language were not reported, however, nor was the language status of the clinicians or whether they could provide SLP service delivery in those languages. Minimal information about racial, ethnic or cultural identity was reported, though 99.1% reported being Australian citizens and less than 1% identifying as being of Aboriginal or Torres Strait Islander descent (Nancarrow et al., 2023). To our knowledge, no information about Australian SLP students has been collected or reported to date.

In the United Kingdom (UK), the Health and Care Professions Council (HCPC) conducted a voluntary survey to Speech-Language Therapists (SLTs) and found that most were white (87%) cisgender (97%) women (95%) (HCPC, 2023). No information about culture or language was reported. The Royal College of Speech Language Therapists (RCSLT) routinely collects and reports data on SLT students in the United Kingdom. Results from the 2020-21 year suggest that the majority were females (95%) and that SLT students were less racially and ethnically diverse than the general student population (RCSLT, 2021a). The most reported racial identities were Caucasian (83%), followed by Asian (9%) Black (3%) Mixed (3%) and other (1%), and most students (7/10) come from households where caregivers or parents hold “intermediate occupations” or “managerial and professional occupations” suggesting they belong to middle or upper-middle-class socio-economic classes (RCSLT, 2021a). Consistent with the survey to clinicians, the survey to students did not examine the cultural or linguistic backgrounds of SLP students in the UK.

Across the United Kingdom, Australia and the United States a clear pattern emerges. Practicing SLPs and SLP students are primarily white, English-speaking females. Prior Canadian surveys suggest a similar trend. In 2021 Speech-Language Audiology Canada (SAC), carried out an Equity Diversity and Inclusion Survey of its members (SAC, 2021). Findings indicate that most SAC members are women (93.4%). The proportion of newcomers in SAC (15.1%) was lower than that in Canada at large (21.9%), as was the proportion of Indigenous members (1.2%) relative to Canadians (6.1%), the proportion of members with racial identities other than Caucasian (13.%) relative to Canadians (22.3%), and the proportion of members who spoke a language other than English or French as their native tongue (11.4%) relative to Canadians (21.1%) (SAC, 2021).

However, no information was reported regarding the nature of the racial or ethnic composition of SAC members (e.g., all racial identities were labelled as ‘people of colour’), their language status, the languages they spoke, or their proficiency to provide service in these languages. Furthermore, this survey was only sent to members of SAC and received only a 22% response rate. Not all Canadian SLPs are required to belong to SAC and so it is possible that results do not reflect SLP demographics across the provinces and territories equally. Furthermore, results are reported for all members, including audiologists and SLPs. No separate data by profession was shared and no information about SLP students was collected.

In Ontario, the most populated province in Canada, the College of Audiologists and Speech-Language Pathologists of Ontario (CASLPO) began collecting demographic data from its members in 2022 through an optional survey embedded into the annual registration renewal process. In 2023, of the estimated 4,600 registered members, 2,861 opted to share their demographic data; a 60% completion rate (CASLPO, 2024). In terms of race, most respondents identified as White/ Caucasian (68%), followed by South-East Asian (15%), South Asian (7%), Black (7%) and other/prefer to not say (8%), while Indigenous populations made up 1% of respondents (CASLPO, 2024). In total, 96% of respondents spoke English, 0.3% spoke French and 3% of respondents reported an ability to speak another language (CASLPO, 2024). There were 101 unique languages represented across the respondents, including English and French. No information was reported regarding language status, or capacity for service in these languages (CASLPO, 2024). Most respondents were areligious (44%) or Christian (33%). No additional data regarding cultural identity was collected or reported. It is promising that CASLPO intends to collect this data on an ongoing basis. However, this represents only a proportion of Ontario-based SLPs and does not reflect all of Canada. In summary, with currently available data, we cannot meaningfully compare the demographic composition of Canadian SLPs or SLP students to the Canadian population at large.

### Rationale for the Current Study

Prior research suggests that SLPs are a relatively homogenous group of white English-speaking females that may not represent the diverse countries where they practice (e.g., ASHA, 2023; CASLPO, 2024; RCSLT, 2023; SAC, 2021). A lack of diversity in the SLP profession is likely to have clinical implications because SLPs assess and provide intervention for communication skills, which are mediated by *language, race,* and *cultur*e. Languages differ in their technical characteristics. They each have unique sound systems (phonology), different labels to describe things or actions (vocabulary) and unique ways of combining parts of words (morphology) and words together to make sentences (syntax). SLPs evaluate and provide intervention for these technical characteristics of the language(s); SAC, 2016). A high-quality assessment involves evaluating a client in all their languages, a task made more challenging when the clinical community lacks linguistic diversity (ASHA, n.d.).

However, it is not only the technical characteristics of a language that impact communication differences across languages and that can be affected by a lack of diversity in the profession. Languages have evolved to be relevant, useful, and specific for the culture and region to which they belong (Steels, 2011). To illustrate the interplay between language and culture, consider a vocabulary assessment where a client is asked to label a picture of broccoli. In English speaking countries like Canada and the United States, we would expect the client to use the English vocabulary term “broccoli” to label this common vegetable. In other places where the vegetable is also common, like Mexico, Portugal, and Germany, it is labelled as “brócoli,” “brócolis” and “Brokkoli” respectively. These cultures are familiar with the vegetable and use a different linguistic term to label it. However, in Gujarati and Urdu, languages spoken in countries like India and Pakistan, broccoli is uncommon and may not be part of the cultural experience of these speakers. A client seeing this image might be unfamiliar with its label, not because of vocabulary difficulties, but because of cultural differences. Beyond semantics, culture can also affect language use or pragmatics, and this can have implications for assessment and intervention. For instance, in Western cultures, it is often expected that an interlocutor will respond directly to questions and make eye contact with their communicative partner when speaking to them. However, in other cultures, it may be perceived as rude or a sign of disrespect to look someone directly in the eye (Unono & Hietanen, 2015). If clinicians are unaware of these cultural communication differences, they run the risk of misinterpreting them as disorders. A lack of diversity and representation of minority communities is known to result in poorer outcomes for clients (e.g., Hersh et al., 2014).

The first steps in increasing diversity are understanding the current composition of the clinical population, and tracking changes over time. Collecting this type of demographic information about all clinicians practicing across Canada would likely only possible with the support and infrastructure of a federal organizing body, such as SAC mandating it, and is beyond the scope of this study. However, given the limited number of clinical education programs nationally, it is feasible to collect demographic information from the clinical SLP students who are currently enrolled in one the twelve Canadian institutions that host a clinical SLP Master’s program. Examining diversity of this group can provide a snapshot into the demographics of the future clinical population; can provide information regarding how representative the future clinical SLP will be of the Canadian population; and can give insights into where our profession may need to focus efforts for change. Canada’s population has always been diverse. However, it has become increasingly so in recent years. Between 2016 and 2021 Canada accepted over 1.3 million newcomers. This change in population has resulted in increased linguistic, racial, and cultural diversity (Statistics Canada, 2022d). As such, SLPs will encounter even more clients from various racial, cultural, and linguistic backgrounds in their practice . Given that communication, a major focus of SLP services, is a linguistically and culturally mediated act, it is critical to examine demographic characteristics of the student SLP population, to determine whether future SLPs are representative of the Canadian population at large in terms of their racial, ethnic, cultural and linguistic identities. Below we define these terms and their connection to communication. Definitions are derived from those used in the Canadian Census.

In this study, *race* is used to refer to shared physical characteristics that are formed through common ancestry. The term visible minority is used to reflect race in the Canadian Census (Statistics Canada, n.d.). Race differs from *ethnicity*, which refers to groups with shared cultural identities (Statistics Canada, n.d.). *Culture* encompasses social customs, beliefs, and knowledge and can influence language. *Language* is the means of communication within a society, be it verbal, written, or signed. The community and culture within which one lives influences what they are exposed to and what language they may have to label items, some of which may not exist in other cultures, and thus other languages.

Within this study, the separation of these concepts allows for a richer understanding of the ways in which they can all interact from the perspectives of SLP service and clientele. For instance, one may belong to an ethnic group and share in its culture, but they may not speak the language shared within that ethnicity; thus, they have cultural knowledge, but not linguistic knowledge. In a similar vein, sex and gender have been separated, as well. *Sex* refers to the sex assigned at birth, typically based on one’s reproductive system and physical characteristics (Statistics Canada, n.d.). *Gender* refers to the gender identity one internally feels and/or the gender they may publicly express in their daily life (Statistics Canada, n.d.). Lastly, *socioeconomic status (SES)* was defined in this study as total household income. Prior to outlining the specific objectives of this survey study, we provide a brief overview of the relevant demographic characteristics of the Canadian population.

#### Languages spoken in Canada

The 2021 Canadian census found that while Canada’s two official *societal languages*, English and French, remained the most spoken, 12.7% (4.6 million) of Canadians reported speaking another language at home (Statistics Canada, 2022i). Four hundred and seventy-four unique languages were reported to be spoken across the provinces and territories. (Statistics Canada, 2022i). The most common heritage languages (i.e., a non-societal language learned by speakers as children, typically in home settings) spoken by Canadians include Mandarin (1.9%), Punjabi (1.4%), Cantonese (1.0%), Spanish (0.9%), and Arabic (0.8%; Statistics Canada, 2022i). The number of speakers of other heritage languages, namely South Asian and African languages has also rapidly increased since the 2016 census. These include Tigrinya (+114%), Malayalam (+129%), Hindi (+66%) and Gujarati (+43%). Finally,72 Indigenous languages were reported to be spoken in Canadian households. Given this linguistic diversity, it is unsurprising that more than 40% of the Canadian population reported being bilingual, and 9% reported being trilingual (Statistics Canada, 2022i).

#### Racial and ethnic identity of Canadians

The proportion of individuals with racial identities other than the Canadian majority has also grown. Approximately 25% of the population identifies as a member of a racialized group or racial minority (2022). This proportion has more than doubled since 2001(13.4%) and more than quintupled since the 1980s (5%). The most common racial identities reported in the 2021 census were South Asian (7.1%), Chinese (4.7%) and Black (4.3%). Each of these groups had a population greater than one million. Other groups that comprised more than 1% of the total population included Filipino (2.6%), Arab (1.9%), Latin American (1.6%) and Southeast Asian (1.1%%; Statistics Canada, 2022b). While many of these individuals were newcomers to Canada, approximately 3/10 were Canadian born (Statistics Canada, 2022g). With newcomers comprising the largest share of the Canadian population in over 150 years (23%), and with population projections estimating that newcomers could comprise between 29-34% of the country by 2041, the country’s racial and ethnic diversity will continue to grow (Statistics Canada, 2022g).

#### Cultural identity of Canadians

Culture is multidimensional and can overlap with race and ethnicity, and with other characteristics like language and religion. How an individual describes their cultural identity can be flexible, change over time, and can depend on their knowledge of their family history, their social environment and even current local and global events. For many their racial/ethnic and cultural identity may be synonymous. For instance, the first author is a fourth generation Canadian, who would report Canadian as both an ethnic and cultural identity. However, for others these identities may differ. In the 2021 census, the most reported cultural identity was Canadian (15.6%), followed by the British Isles including English (14.6%), Irish (12.1%) and Scottish (12.1%) (Statistics Canada, 2022e). French and French-Canadian cultural identities were reported by 11% and 2% of Canadians respectively (Statistics Canada, 2022e). Additionally, more than one million people reported their cultural identity as German (3 million), Italian (1.5 million), Ukrainian (1.3 million), Chinese (1.7 million) and Indian (1.3 million) (Statistics Canada, 2022e). There are also three groups that are considered as Indigenous peoples in Canada. These are Inuit, Métis and First Nations (Statistics Canada, 2022h). Collectively, more than 2.2 million individuals reported Indigenous ancestry or identity in the 2021 Census, approximately 5% of the Canadian population and a 9.4% increase from 2016 (Statistics Canada, 2022h).

#### Goal of the Current Study

In this study we collected demographic information about Canadian clinical SLP students currently enrolled in one the country’s twelve accredited institutions. To provide a detailed picture of the characteristics of current SLP students, we surveyed students about their age, sex and gender; their racial, cultural, and ethnic identities; their language status, the languages they speak, their proficiency for service in these languages; and their socioeconomic status.

## Method

### Ethics

Procedures as outlined in the study protocol received approval by the University of Toronto’s Health Sciences Research Ethics Board (Protocol #45795).

### Participants

To be eligible to participate in this survey, individuals must have been enrolled as a student in one of the twelve accredited Canadian Speech Language Pathology Master’s programs, during the 2023-24 academic year.

### Recruitment

Recruitment was carried out in the spring of 2024, between March and June. To increase response rate, the research team identified course instructors teaching Year 1, Year 2 or Year 3 SLP students in spring 2024 (N=25). We contacted these instructors via their university emails in either English or French, depending on the language of instruction of the program. We provided a description of the study, and our research aims and requested a brief virtual meeting. In these meetings, we addressed any questions instructors might have about the study protocol, the ethics approval, the data collected and/or the purpose for the study. Subsequently, we shared the ethics-approved infographic/slide with survey link to the instructors in either English or French via email. We requested that course instructors share the graphic and survey link with their Year 1, 2 or 3 SLP students during class time. In in our experience, students are more likely to complete a survey if they learn about it from a course instructor and are provided with in class time to complete it, than if they are sent a survey link via email and expected to complete it on their own time. A higher response rate is crucial for a survey of this nature as it better reflects the true demographics of the population of interest. Despite our best efforts, in some instances, the survey could not be shared with students in class (e.g., the Year 3 students at Dalhousie were finished with coursework at the time of data collection and were on placement). For 7 of the 25 cohorts, the survey link was sent to students via email or via course online platform message by a course instructor or program director. Course instructors and professors also reported the number of students in their cohorts to the research team. In total, there were 987 SLP students reported to be currently enrolled in either their first, second or third year of an accredited master’s program in the twelve Canadian institutions in Spring of 2024 who were eligible to be recruited for participation in this study.

### Measures

Study measures were developed and hosted on REDCap, a secure web application used by the University of Toronto for survey dissemination (Harris et al., 2009). When accessing the survey link, students were first prompted to confirm their eligibility to participate by reporting whether they were enrolled in a clinical Speech-Language Pathology Master’s program at a Canadian institution. If eligible, they were then directed to the informed consent page. Eligible participants were provided information about the study, risks and benefits, accessing research results, the type of data to be collected and how and where it would be stored, and were encouraged to contact the research team with any questions. Participants documented their consent by confirming “Yes”, rather than signing, as the survey was anonymous.

Participants who consented were automatically directed to the survey. To further increase response rate, we elected to keep the survey short, and to prioritize collecting information about demographic characteristics of greatest interest and relevance to the evaluation and intervention of communication difficulties (e.g., prioritizing questions about language, racial, and cultural identity over those related to prior education, work experience or sexual orientation). Wherever possible, we also elected to use terminology and question framing that was consistent with the language and structure of the Statistics Canada 2021 Census to ensure that the data we collected about Canadian SLP students could be easily compared to the broader Canadian population.

The demographic survey had between 17-24 questions (depending on the number of languages spoken by a participant) and took approximately 5-10 minutes to complete. General survey questions addressed respondents’ institute of study, year of graduation, age, sex, and gender identity. Language questions queried language status (e.g., number of languages spoken as a measure of mono-, bi-or multilingualism) and languages spoken (e.g., official societal languages such as French and English vs. heritage languages such as Tamil or Mandarin). Students were also asked to self-rate whether they had the capacity to provide clinical services in their spoken languages (i.e., by indicating yes or no for each language spoken). In defining their racial and ethnic identity, participants could select multiple identities from the list of 450 identities developed by Statistics Canada. For cultural identity, participants could type in any other identities or information that formed part of their culture. Questions about SES included selecting from a list of income ranges that best matched their estimated total household income, and reporting how many individuals that total income supported. At the end of the survey, participants had the option to leave questions or comments, and to provide their email if they wanted to be informed about study outcomes. For all survey questions, students had the opportunity to either select “Prefer to not disclose” or type “N/A” if they did not wish to disclose the information. All study materials were available in both English and French. A copy of the demographic survey can be found in the Appendix.

### Analysis

Following data collection, anonymized data was extracted from REDCap and transferred to an excel file in preparation for analysis. Given that the primary aim of this study was to describe the demographic characteristics of Canadian SLP students, descriptive statics were used for analysis. For continuous data, means and ranges were calculated and reported (e.g., age). For nominal data (e.g., languages spoken) totals were counted and percentages of the whole were calculated and reported. For instance, 60.5% (319/525) of surveyed students reported their first language as English.

## Results

Across the twelve institutions 525/987 eligible students completed the survey, for a total response rate of 53%. Response rates ranged from 29% (Université de Laval) to 86% (McGill University). For a detailed summary of response rates, see Table 1 in the Appendix.

### Age, Sex and Gender

Survey respondents were first asked to provide their age, sex and gender identity. The average age of current Canadian SLP students was 25 (range 22-48). The vast majority of SLP students (96%) were female (N=504) while only 3.6% were male (N=19). Less than 1% did not disclose their sex (N=2). A small percentage SLP students (N=11, 2%) did not report their gender identity. Of those who did, 96% were cisgender (N=505) and 1.3% identified as either “non-binary” (N=5), “genderqueer” (N=1) or “trans” (N=1). Two students reported their sexual orientation (e.g., heterosexual or straight) rather than gender in response to the survey prompt and therefore data about their gender identity was not captured. In summary, the majority of Canadian SLP students are young, cisgender females.

### Language Status and Languages Spoken

Survey respondents were asked how many languages they spoke, what those languages were and whether they felt capable of providing SLP services in the languages. Of the 525 respondents, only a third 33.9% were monolinguals (N=178), while the largest proportion, 47%, were bilinguals (N=247) and the smallest percentage, 19%, were multilinguals (N=100).

#### First Language

The majority of respondents reported one of Canada’s two official societal languages, either English (N=319, 61%) or French (N=148, 28%) as their first or native language. The most common heritage languages reported by respondents included Arabic (N=8, 1.5%), Spanish (N=8, 1.5%), Urdu (N=5, 0.9%), Cantonese (N=4, 0.8%) and Turkish (N=4, 0.8%). In Figure 1 below, each first language spoken is represented by a unique circle. The circles are proportional in size to the percentage of speakers who reported that language as their first/native language. Societal languages of English and French are depicted in warm tones, while heritage languages are depicted in cool tones.

**Figure 1.**
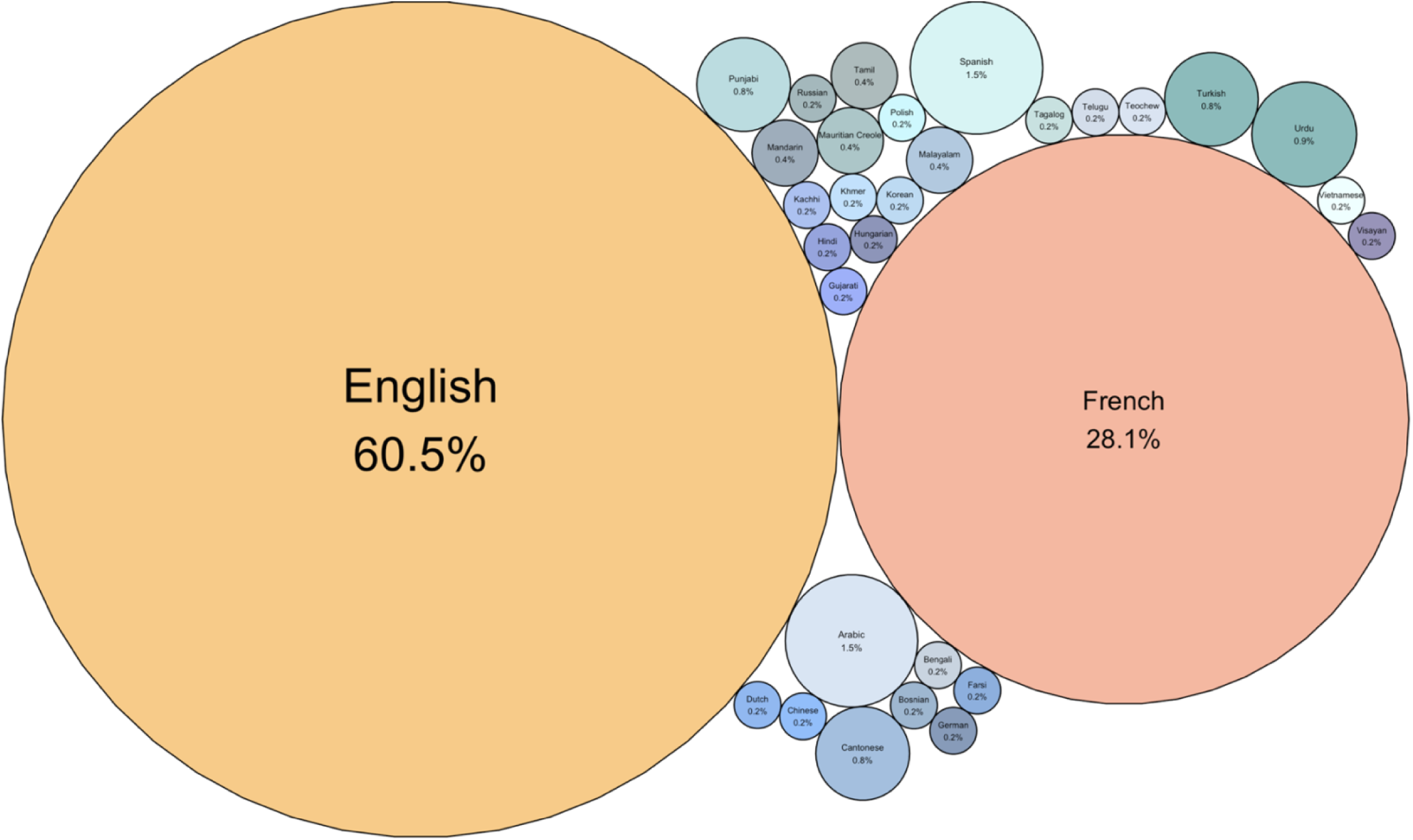
First languages spoken by Canadian SLP students **Note.** N=525. In this figure, each language is represented as a unique circle. The circles are proportional in size to the percentage of students who reported that language as their first/native language. Societal languages are represented in warm tones and heritage languages are represented in cool tones.

#### Second Language

Of the respondents, 347 were bi or multilingual and spoke a second language. Canada’s two official languages were also the most commonly reported second languages by Canadian SLP students. Half (N=173, 50%) of all bi or multilingual speakers reported English as their second language, and a third (N=114, 33%) reported French as their second language. Beyond English and French, the most commonly spoken second languages were Spanish (N=7, 2%), Arabic (N=6, 1.7%), Cantonese (N=5, 1.4%), Portuguese (N=4, 1.1%), Punjabi (N=4, 1.1%), Tamil (N=4, 1.1%), Mandarin (N=3, 0.8%) and Urdu (N=3, 0.8%). This suggests that the most common bilingual profile among Canadian SLP students is English/French. Refer to Figure 2 below, which depicts the percentage of respondents’ reported second languages.

**Figure 2.**
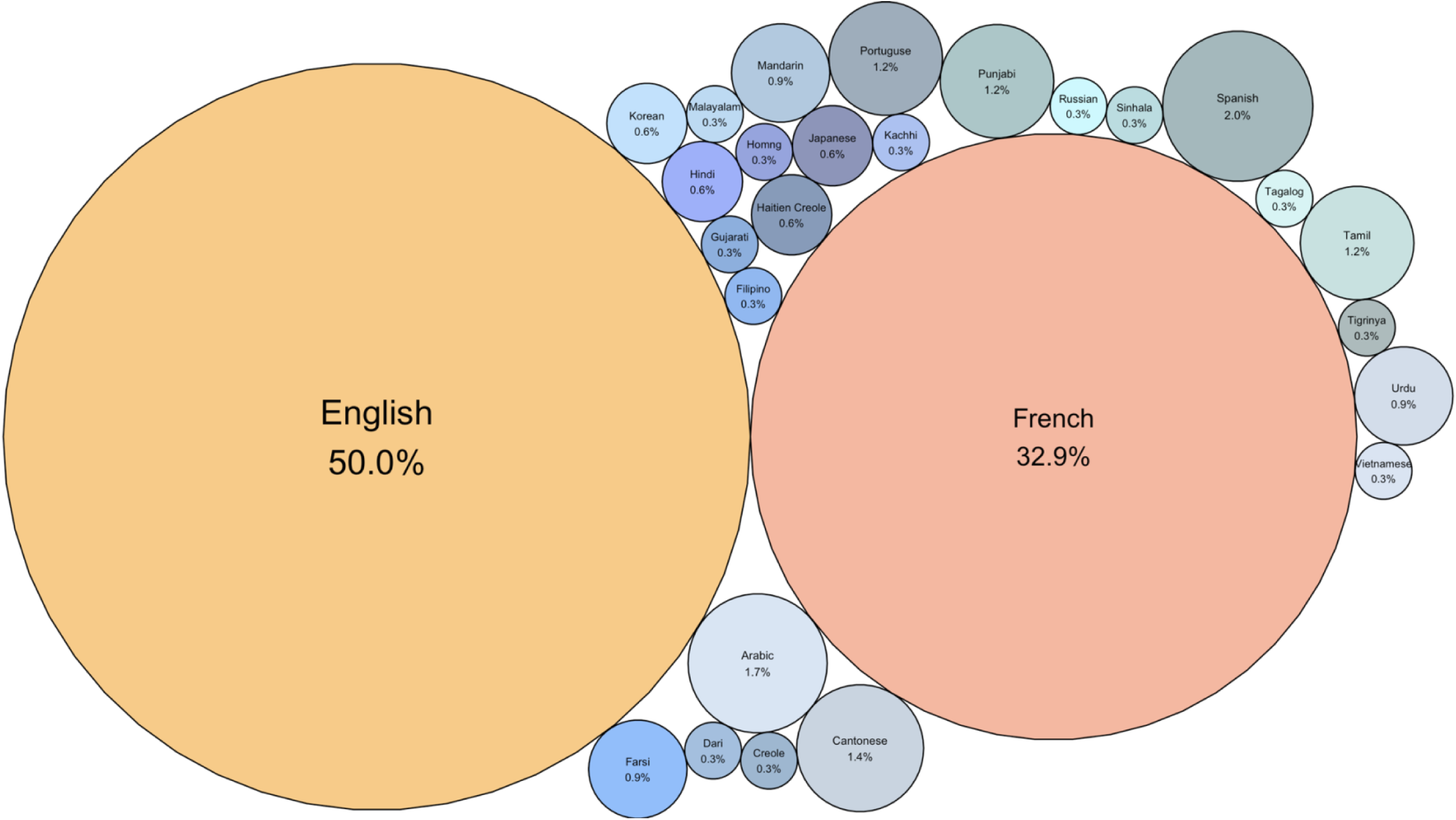
Second languages spoken by Canadian SLP students **Note.** N=347. In this figure, each language is represented as a unique circle. The circles are proportional in size to the percentage of students who reported that language as their second language. Societal languages are represented in warm tones and heritage languages are represented in cool tones.

#### Third Language

One hundred SLP students were multilingual and spoke three or more languages. Spanish was the most commonly reported third language (24%), followed by French (20%), English (15%), Hindi (5%), Italian (5%), and Greek (4%). Some students also spoke a fourth and fifth language. These languages are not represented in Figure 6 below, but are Spanish (N=1), Polish (N=1) and Zaghawa(N=1). Across all languages, there were 48 unique languages spoken by Canadian SLP students. Refer to Figure 3 which depicts the percentage of respondents’ reported third languages.

**Figure 3.**
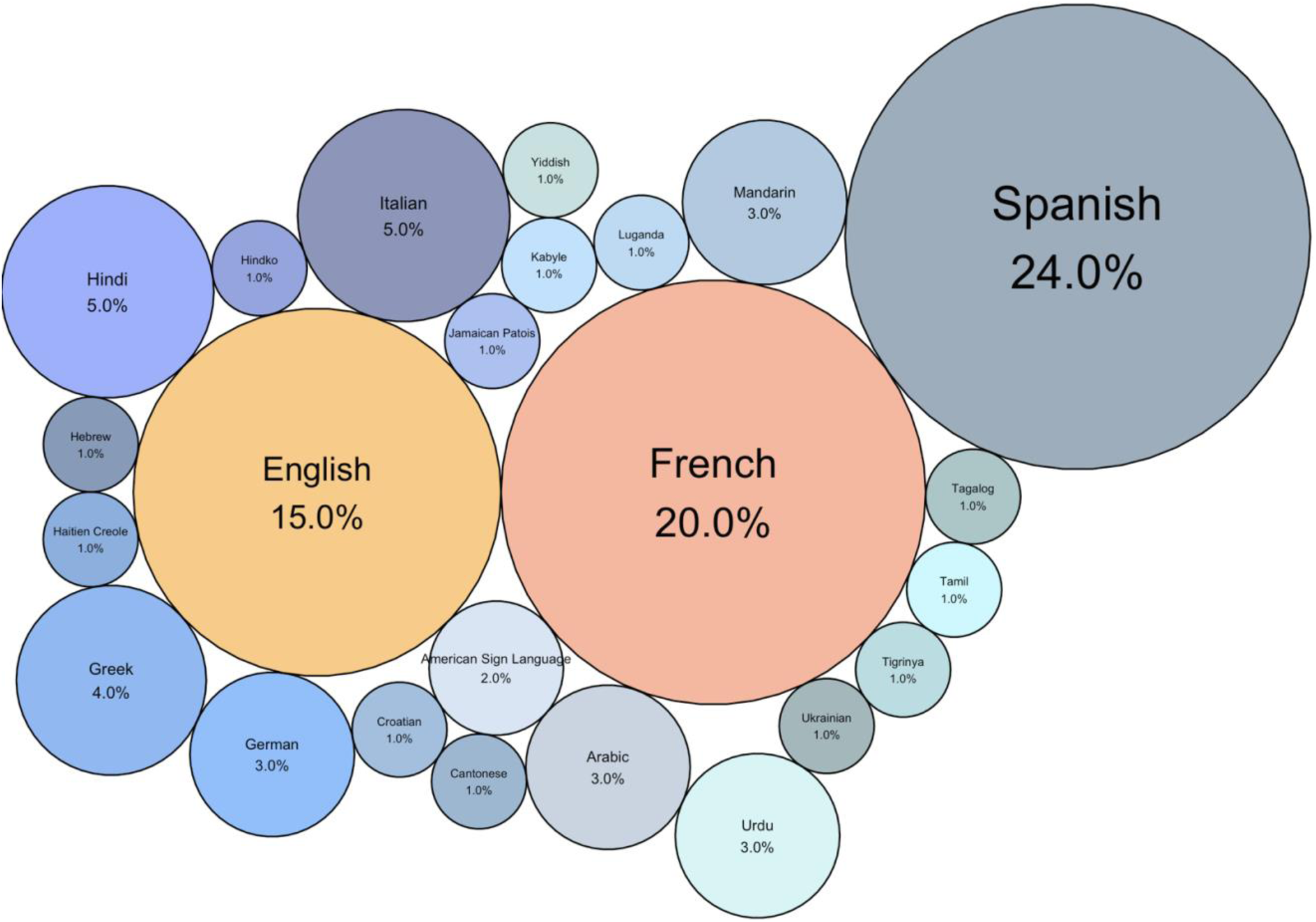
Third languages spoken by Canadian SLP Students **Note.** N=100. In this figure, each language is represented as a unique circle. The circles are proportional in size to the percentage of students who reported that language as their first/native language. Societal languages are represented in warm tones and heritage languages are represented in cool tones.

### Capacity for Service Delivery Across Languages

Surveyed SLP students were then asked to report whether they felt capable of providing SLP services in their spoken languages. Nearly all of those who reported English (99.1%) or French (99.3%) as their first language indicated that they could provide SLP service in these languages. However, only 76.3% of those who reported another language as their first felt capable of providing services in it. A similar pattern emerged for capacity to provide service in a second language. For those for whom English was their second language, 63% felt capable of providing service in this language, while 46.4% felt capable of providing service in their second language of French. Only 37.3% of respondents who spoke another language as their second felt capable of providing service in that language. This trend continued for capacity to provide service in a third language. More than two thirds (66.7%) of those who spoke English as their third language and more than a quarter (27.8%) of those who spoke French as their third language reported feeling capable of delivering SLP service in these languages. Only 20% of survey respondents who spoke another language besides English or French as their third language reported feeling capable of providing service in that language.

Overall, results suggest that SLP students are less likely to report feeling capable of providing services in their second and third languages and are also less likely to report feeling capable of providing service in languages other than English and French regardless of whether it is their first, second or third language. The stacked bar graphs in Figure 4 below depict the percentage of SLP students who reported whether they did or did not self-rate as capable of providing service in English, French or other languages when these languages were their first, second or third language.

**Figure 4.**
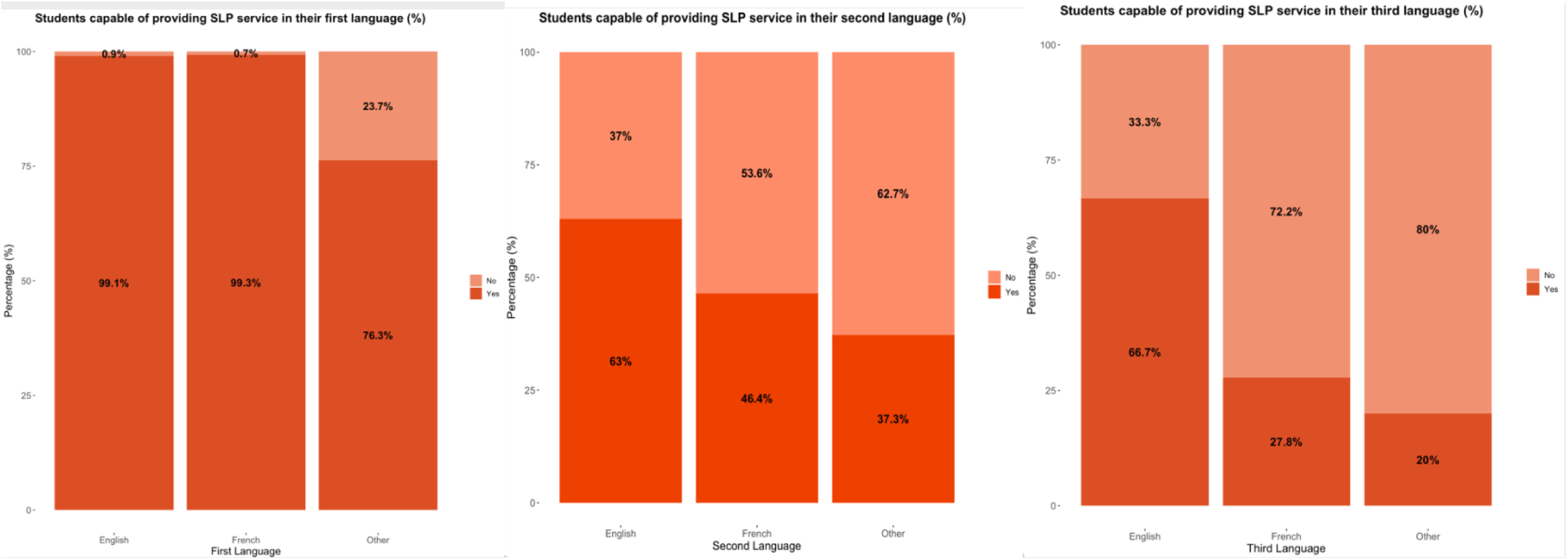
Students capable of providing SLP services in their first, second and third languages **Note.** The stacked bar graphs above depict the percentage of students who self-rated as either capable (“yes”) or not capable (“no”) of providing SLP services in Societal languages (English and French) vs. Heritage languages (Other). From left to right the three graphs depict this capacity when these societal and heritage languages are the students’ first (N=525), second (N=347) and third (N=100) language respectively. Percentages are used rather than counts to reflect proportion and to allow for comparison across first, second and third languages.

Of the 48 languages reported to be spoken by SLPs, SLPs students reported feeling capable of providing service in 33 (not including English or French) This represents 69% of the total number of unique languages spoken. Many of these languages were spoken by small numbers of students. Of the 525 students surveyed, only 80 (15%) reported being capable of providing service in a language other than English or French. The most common heritage languages SLP students felt capable of providing service in were Spanish, Punjabi, Arabic, Urdu, Cantonese, Hindi and Tamil. Please see Figure 5 for a breakdown of the number of students who reported capacity to provide SLP service delivery in languages other than English and French.

**Figure 5.**
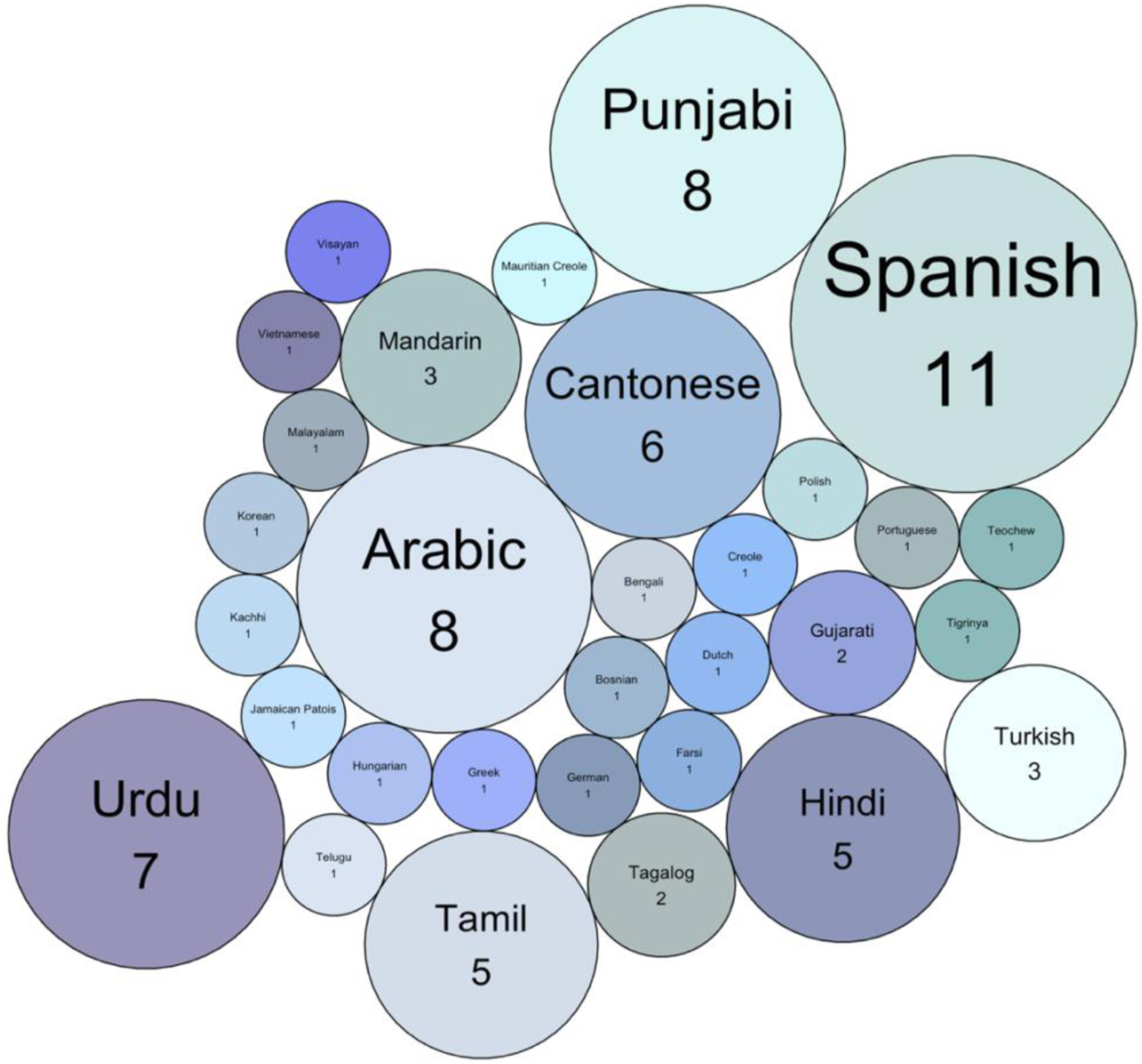
Languages in which SLP students can provide services **Note.** N=80. In this figure, each heritage language is represented as a unique circle. The circles are proportional in size to the number of students who reported that language as their first/native language.

**Figure 6.**
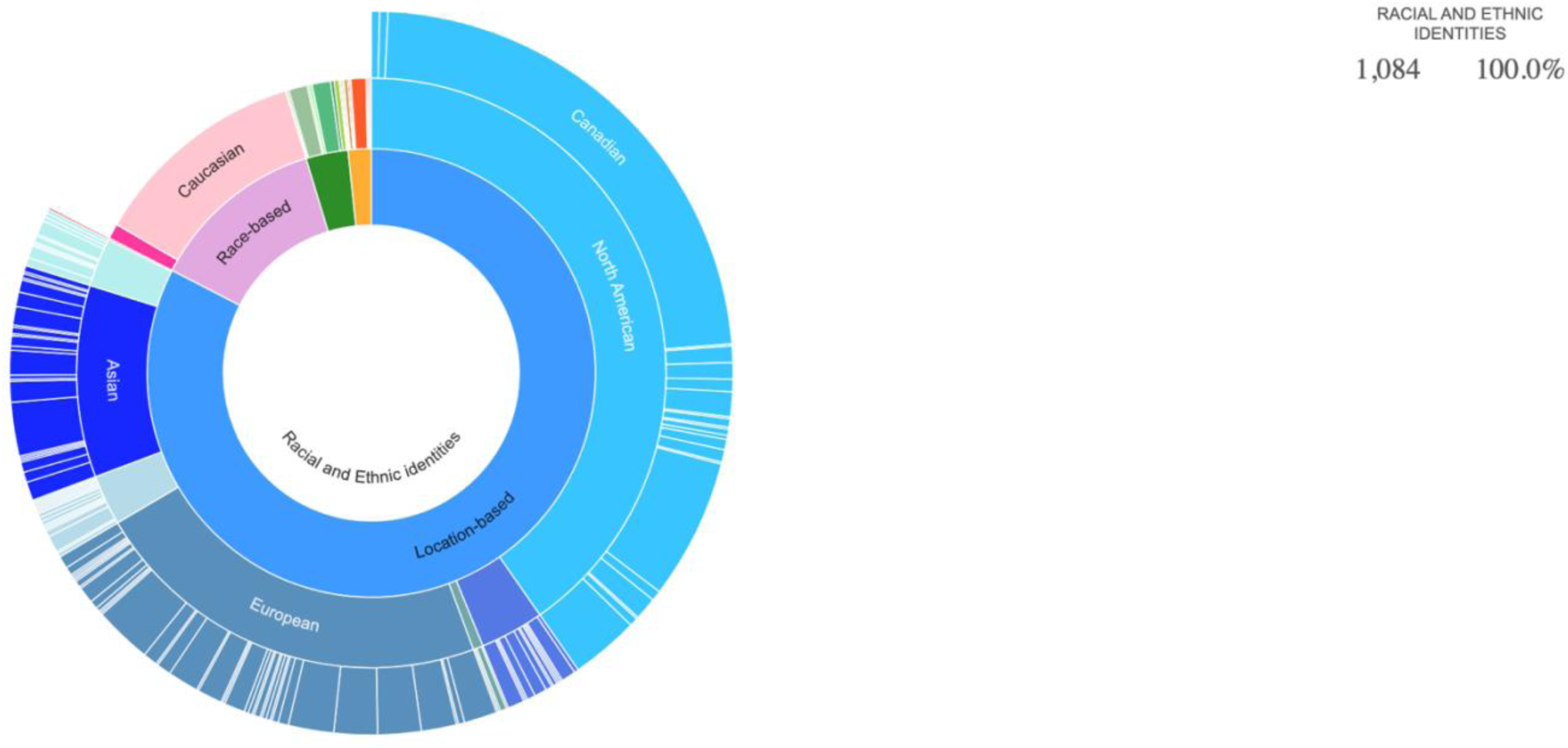
Racial and ethnic identities reported by Canadian SLP Students **Note.** The above is a static image of an interactive sunburst plot, which represents hierarchical levels of responses. In the inner most circle, all 1084 responses reflecting racial and ethnic identities are represented. In the next level of circle, responses are segmented into location, race, religious and Indigenous-based identities. In the third level, the different subcategories of location (e.g., continents), race (e.g., Black), religion (e.g., Christian) and Indigenous identities (e.g., Métis) are reflected. Finally, in level 4, the unique locations reported by all students are reflected within each continent. Wedge sizes correspond to the proportion of respondents who selected the (category) of identity. The interactive plot can be viewed at this link: https://rpubs.com/EmilyKW/1221010 When viewing the interactive plot, clicking a segment will populate information about the number and percentage of respondents who selected that identity.

### Race and ethnicity

Survey respondents were asked to report racial and ethnic identities with which they identified. The list of identities was copied from the list used in the Statistics Canada 2021 Census and included 450 unique options (Statistics Canada, 2022b;2022e). Racial and ethnic identities were characterized in different ways. Some racial and ethnic identities related primarily to a location, either a continent, country, province or territory, or geographic region (e.g., “North American”, “Canadian”, “Québec, “Middle Eastern”). Other options from the list were tied to religious or spiritual beliefs (e.g., “Christian”, “Muslim”, “Jewish”). Fewer identities related to race as skin colour (e.g., “White”, “Black”). Finally, some identities described Indigenous groups (e.g., “Cree”, “First Nations”, “Haida”).

SLP Students could select as many options from the list as they felt necessary to represent their racial and ethnic identity. A total of 1084 identities were reported by the 525 respondents, for a mean of 2 identities per student. The range of identities reported per student was 1-12. Across these 1084 responses, 151unique identities of the 450 options were reflected. Most students identified themselves by location (N=895, 82.6%) and indicated racial and ethnic origins from North America (N=437, 40.3%) or Europe (N=240, 22.1%). More than 10% of students indicated Asian racial ethnic origins (N=114), but very few indicated Central American (N=33, 3.5%), African (N=30, 2.8%), Middle Eastern (N=29, 2.7%) or South American (N=6, 0.6%) identities. Some students identified themselves by race (N=135, 12.7%). Of these, most (N=129, 11.9%) selected Caucasian (White) while very few selected Black (N=9, 0.8%). A small proportion of student SLPs identified themselves by their religion (N=33, 3.0%). The most reported religions were Christian (N=11, 1.0%) and Jewish (N=11, 1.0%). There were only 18 responses that reflected Indigenous identities from the surveyed students (1.7%). Of those, the most common Indigenous identity was Métis (N=9, 0.8%). Additional information about the number of responses for each location, race, religious and Indigenous-based identity reported can be found in Figure 6 below. This sunburst plot is hierarchical and interactive. Readers can first select the overarching racial/ethnic category of interest (e.g., location-based or race-based), and then select further within that category (e.g., North American or Black). The count and percentage in the top right corner will automatically reflect the selected categories.

### Culture

The demographic survey also included a question where students were asked to report their cultural identity. As stated, culture *can* overlap with racial and ethnic identity. However, one’s racial and ethnic identity *may not always* be equivalent to their culture (Statistics Canada, 2021). For instance, one respondent indicated in a comment on the survey that while she identified racially and ethnically as Chinese, she felt her cultural identity was better represented as Canadian, as she had been born, raised and schooled in Canada. This survey question was open-ended, and participants were encouraged to describe their cultural identity in whatever way best reflected them.

Of the 525 survey respondents, 470 answered this question (90%). A total of 768 cultural identities were reported. Respondents reported between 1 and 5 cultural identities each, with an average of 1.5 identities listed per participant, and 121 unique identities reported across all responses. Much like for racial and ethnic identity, cultural identities could be broadly categorized as location-based (e.g., “Canadian”, “German”), religion-based (e.g., “Atheist”, “Baha’i”), race-based (e.g., “Black”, “Caucasian”), Indigenous-based (e.g., “Aboriginal”, “Cree”). There were several other cultural identities that did not fit neatly into these categories. These include “Conservative,”, “Desi”, “Latin American”, “Latin/Latinx”, and “Queer”.

As with racial and ethnic identity, most students identified themselves by location (N=654, 85.2%). A larger proportion of SLP student respondents identified as culturally Canadian (N=363, 47.4%), compared to racially or ethnically Canadian (N=249, 23 %). Relative to racial/ethnic identity, fewer students reported race-based (N=6, 0.8%), or Indigenous-based (N=6, 0.8%) cultural identities. More than double the number of students reported religion as part of their cultural identity (N=74, 9.7%) compared to their racial or ethnic identity (N=33, 3.0%). The most reported religious cultural identities were Muslim (N=14,1.8%) Christian (N=11, 1.4%), Catholic (N=10, 1.3%) and Jewish (N=10, 1.3%). Several cultural identities were mentioned that were not reflected in racial and ethnic identity data. Of these, the largest was “Desi” (N=18, 2.3%) a group that describes those with familial origins in India, Pakistan or Bangladesh who reside outside of those countries (Charles, 2018). Some students felt their identities were best reflected by their political values, as in the student who reported “Conservative” as their cultural identity. Others felt their sexual orientation and/or gender identity best reflected their culture, as in the student who reported “Queer” as their cultural identity. For a comprehensive breakdown of the cultural identities reported by the surveyed Canadian SLP students, please refer to Figure 7 below. This sunburst plot is hierarchical and interactive. Readers can first select the overarching cultural identity category of interest (e.g., religion-based), and then select further within that category (e.g., Muslim). The count and percentage in the top right corner will automatically reflect the selected categories

**Figure 7.**
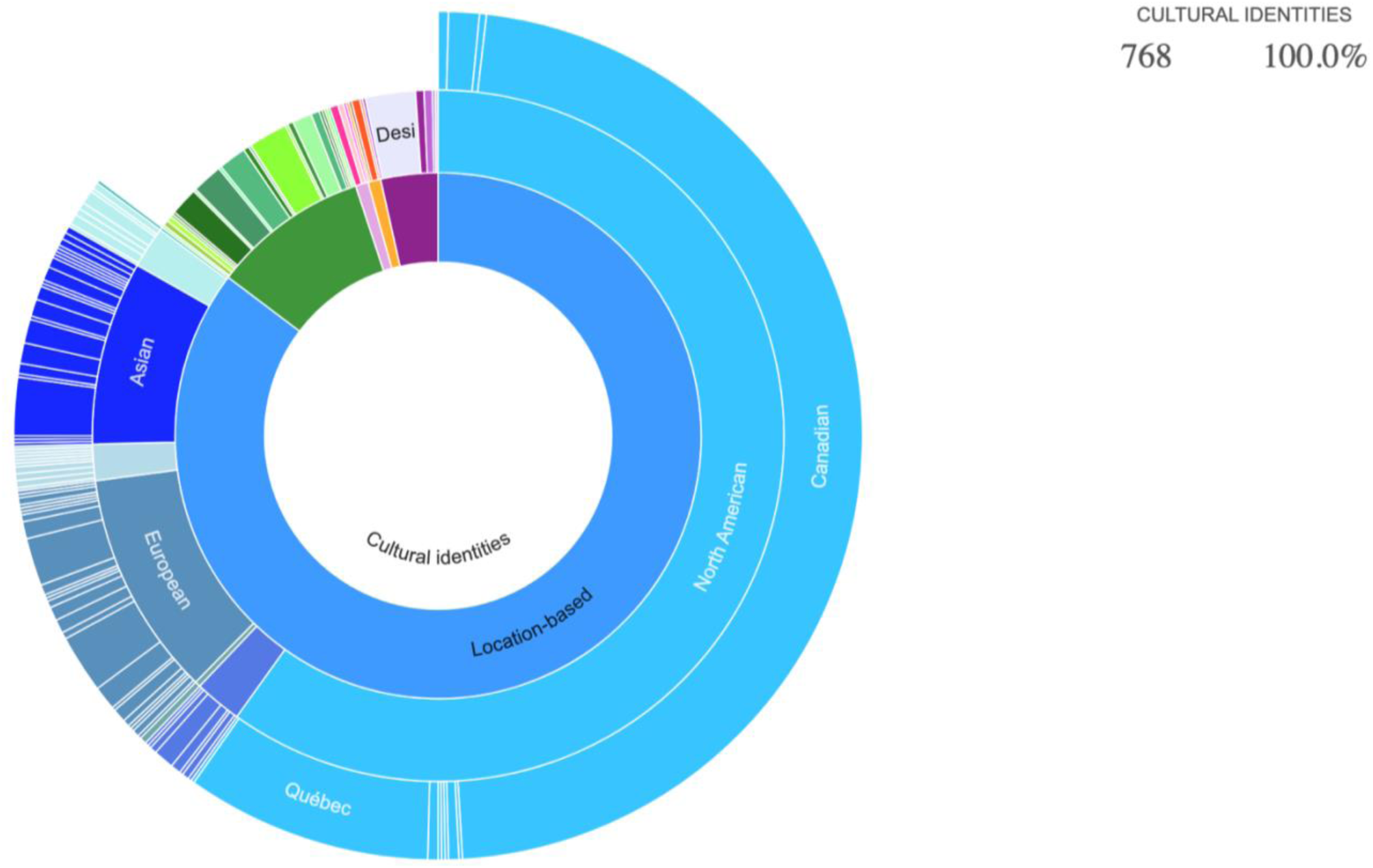
Cultural identities reported by Canadian SLP Students **Note.** The above is a static image of an interactive sunburst plot, which represents hierarchical levels of responses. In the inner most circle, all 768 responses reflecting cultural identities are represented. In the next level of circle, responses are segmented into location, race, religious, Indigenous-based and other identities. In the third level, the different subcategories of location (e.g., continents), race, religion and Indigenous identities are reflected. Finally, in level 4, the unique locations reported by all students are reflected within each continent. Wedge sizes correspond to the proportion of respondents who selected the (category) of identity. The interactive plot can be viewed at this link: https://rpubs.com/EmilyKW/1221013 When viewing the interactive plot, clicking a segment will populate information about the number and percentage of respondents who selected that identity.

**Figure 8.**
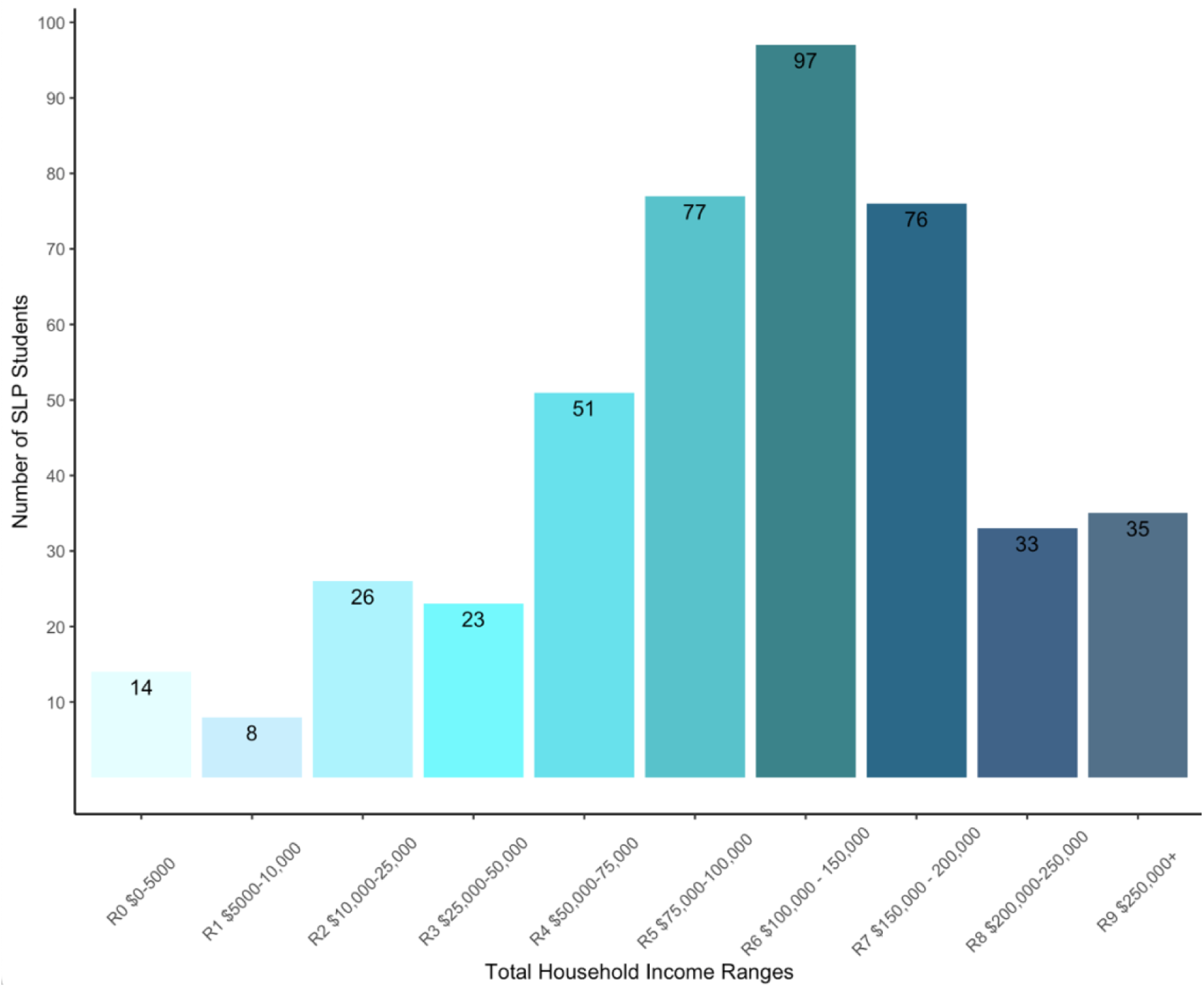
Estimated total household income of Canadian SLP students **Note.** N= 440. This bar graph depicts the number of students who reported estimated total household incomes within each of the 10 income ranges.

### Socioeconomic status

Surveyed SLP students were asked to estimate their total household income and report the number of individuals supported by this income as an indicator of their socioeconomic status (SES). Each student has a unique situation, some live at home with parents or caregivers and are supported by them, others live independently and have taken out student loans or work part time, others may be mature students who have children of their own to support. Students selected from 10 income range options that aligned with the ranges used in the Statistics Canada Census 2021 (Statistics Canada, 2023). The lowest range represented those earning less than $5000.00 per year and the highest being those whose total yearly household incomes were greater than $250,000.00. The highest proportion, and nearly a quarter, of students (N=97, 22%) reported a total household income of $100,000.00 - $150,000.00, followed closely by estimated household incomes in the range of $75,000.00 - $100,000.00 (N=77, 18%) and in the range of $150,000.00-$200,000.00 (N=76, 17%). There were several students who elected to not disclose their household income (N=85, 16%). More than half (N=241, 55%) of those who disclosed their total household incomes reported incomes greater than $100,000.00. In terms of the number of individuals supported by total household income, students reported a range of 1-11 persons, with an average of 3.5 individuals across respondents. Some students did not disclose the number of individuals their total income supported (N=56, 11%). See Figure 12 below for the numbers of students who reported total household incomes in each range.

## Discussion

We conducted a nationwide survey of Canadian speech-language pathology (SLP) students enrolled in one of the twelve accredited clinical Master’s programs in the spring of 2024 to provide a snapshot of the future of the clinical profession. As opposed to previous similar surveys, we reported comprehensive demographic information, including sex, gender, language status, languages spoken, racial, ethnic and cultural identities, and the socioeconomic status of students. Compared to results of large-scale demographic studies of practicing clinicians in Canada (SAC, 2021), and the United States (ASHA, 2023), the Canadian SLP student population is trending toward greater diversity on may factors including bi and multilingual representation, nonbinary and trans representation, and increased racial, ethnic and cultural diversity. However, when compared to the Canadian context, there are metrics where the diversity of the Canadian population is not reflected in the current SLP student cohort. This includes representation of speakers of Canada’s most spoken heritage languages, rates of male, black and Indigenous SLP students, and representation of students from lower SES backgrounds. Below, we first discuss some limitations of our survey to provide a fair context for data interpretation. We then elaborate on how our results compare to the demographics of the broader Canadian population and consider the implications of these findings for future clinicians and the clients they will serve. Finally, we provide recommendations for further diversification of the profession and future research.

### Limitations

While every effort was made to recruit as many Canadian SLP student participants as possible, we ultimately had a response rate of 53%. While not as high as we had hoped, this response rate greatly exceeds SAC’s equity diversity inclusion 22% survey response rate and is on par with CASLPO’s diversity survey 60% response rate from 2022 (SAC, 2021; CASLPO, 2024). Given this, and that we received responses from all cohorts in all Canadian institutions, we feel that the results are broadly representative of the Canadian SLP student population in 2024.

A second limitation of this study pertains to the phrasing used in the question asking SLP students to estimate their total household income and the number of individuals supported by it. Some SLP students commented that they were unsure how to answer this question if they were living alone but had some financial support from caregivers, or if they were living with roommates and sharing some but not all expenses. However, comments of this nature were limited to a few respondents and likely do not impact results regarding total estimated incomes.

### Language

Findings indicate that 47% (N=247) of Canadian SLP students were bilingual and an additional 19% (N=100) were multilingual. In Canada at large, 41.2% of the population is bilingual, and 9% is multilingual (Statistics Canada, 2022i). These findings suggest that rates of bi and multilingualism are slightly higher in the Canadian SLP student population than in the country at large, a trend in a positive direction. However, it is important to not just consider whether clinicians are bi- and multilingual, but also what languages they speak, whether they can use these languages clinically.

Of the 525 surveyed respondents, 507 (96%) spoke English as either their first (N=319), second (N=173) or third language (N=15), and of those, 435 (86%) self-rated as capable of performing clinical services in English. The proportion of SLP students with English as their mother tongue (60.5%) is higher than in Canada at large (54.9%; Statistics Canada, 2022i). The next most spoken language among respondents was French. More than half of the survey respondents (54%, N=282) spoke French as their first (N=148), second (N=114) or third (N=20) language. Fewer respondents self-rated as being able to provide service in French (73%, N=206,). The proportion of SLP students with French as their mother tongue (28.1%) is also considerably higher than in Canada at large (19.6%; Statistics Canada, 2022i). Finally, more than a third of respondents (34%, N=183) spoke a language other than English or French as their first (N=58), second (N=60), or third language (N=65). The proportion of SLP students with a language other than English or French as their mother tongue (11%) is considerably lower than in Canada at large (21%; Statistics Canada, 2022i).

A potential reason for this discrepancy may be that there are barriers for individuals whose native language is not English or French when applying to clinical master’s programs in Speech-Language Pathology. In Canada, 10/12 accredited programs require that “non-native” English or French speaking applicants, and those born outside of Canada, provide proof of English or French proficiency by meeting a threshold score on standardized assessment like the Test of English as a Foreign Language (TOEFL; University of Toronto, n.d.a.). These tests can take several hours to complete and are costly (Educational Testing Service, n.d.) ^1^.

A recent survey of American undergraduate students interested in pursuing SLP found that 32% of students whose first language was not English and who perceived themselves as having accents thought that their accent might present a potential barrier in working in SLP (Fuse, 2018). These prospective students worried that their accent might affect their admission to a program (13%) or their ability to be successful in their future clinical practice (11%; Fuse, 2018). Future research should investigate whether language proficiency requirements discourage potential non-native English and French speaking students from applying and the impact this might have on the linguistic diversity of SLP graduate cohorts. This is particularly important to consider given that evidence supporting language tests’ ability to predict undergraduate and graduate school performance is mixed at best (e.g., Simner & Mitchell, 2007). For instance, one American study found that performance on the TOEFL accounted for only 3% of the variance in graduate student GPA (Cho & Bridgeman, 2012).

In terms of the diversity of languages spoken, there were 48 unique languages reported by survey respondents across first, second and third+ languages. Besides English and French, the most common languages spoken as first, second or third languages by SLP student respondents were Spanish (N=39, 7.5%,), Arabic (N=17, 3.2%,), Urdu (N=11, 2.1%,), Cantonese (N=10, 1.9%), Mandarin (N=8, 1.5%) and Punjabi (N=8, 1.5%,). These results are mostly reflective of the Canadian population at large. Though census data revealed that over 200 mother tongues are spoken across Canada, English and French remain the most spoken languages. These are followed by Mandarin, Punjabi, Cantonese and Spanish (Statistics Canada, 2022i).

However, just because a student had capacity in a language, did not necessarily mean they felt qualified or skilled to engage in clinical service delivery in that language. In fact, only 15% (N=80) of surveyed SLP students self-rated as capable of providing services in languages other than English or French. Among those languages, only 2% of those surveyed reported that they could provide services in Spanish (N=11), 1.5% in Arabic (N=8),1.5% in Punjabi (N=8), 1.3% in Urdu (N=7) and 1.1% in Cantonese (N=6). Notably, only 3 SLP students indicated they would be able to provide service in Mandarin (0.5%). This is discouraging as Mandarin is Canada’s next most spoken language following English and French, with more than half a million individuals speaking it at home (1.9% of the population) (Statistics Canada, 2022i).

There are several reasons why even those participants who speak a language besides English or French might not feel capable of providing clinical service in it. First, all of Canada’s programs are hosted in English or French and typically focus on the manifestation of speech and language difficulties and disorders in one of these languages. This is logical, as these are the two official societal languages used in schools and healthcare settings and spoken by most Canadians (Statistics Canada, 2022i). However, more than 200 languages are spoken in Canada, and more than 40% of the Canadian population is bilingual, meaning future clinicians are highly likely to also encounter speakers of languages other than English or French in their practice (Statistics Canada, 2022i). In fact, an essential competency for SLP students outlined in the competency profile is the capacity to “*incorporate knowledge of, and respond to, the unique needs of linguistically, sexually and culturally diverse populations into practice*” (Canadian Alliance of Audiology and Speech-Language Pathology Regulators, 2018). Similar competencies are required by SLP students in other countries with high rates of linguistic and cultural diversity (e.g., ASHA, 2017.; RCSLT, 2021b).

There are two primary means of developing student competence for practice with culturally and linguistically diverse (CLD) populations in SLP graduate programs. The first is curricular infusion, where CLD issues relevant to the topic are embedded within domain-specific coursework (e.g., teaching how to modify a speech assessment with a bilingual child in a speech sound disorders course). The second is via dedicated CLD courses where domain-general issues in CLD practice (e.g., developing cultural humility or working with interpreters) are addressed in a standalone course (Higby et al., 2024). It has been suggested that a combination of both infusion and dedicated CLD course(s) is most effective for preparing graduate students (Bradshaw & Randolph, 2021). A review of the course lists provided on the websites of Canadian SLP programs suggest only two Canadian institutions (17%) provide a foundational course dedicated to issues in CLD practice. These are (i) the University of Alberta, (*Anti-racism, Diversity and Equity in SLP practice;* University of Alberta, n.d.) and (ii) the University of British Columbia, (*Approaches to Audiology & SLP for People of First Nations, Metis or Inuit Heritage;* University of British Columbia, n.d.). It plausible to assume then that most Canadian institutions use primarily an infusion approach to integrating CLD issues into their programs. Interestingly, a somewhat different trend is observed in the United States. A recent survey of 565 instructors at SLP programs across the United States found that 38% of institutions used a curricular infusion approach alone (Higby et al., 2024). This was down from 56% in 2008 (Stockman et al., 2008). Conversely, 36% used a combination of infusion and dedicated CLD courses (Higby et al., 2024). This was up from 31% in 2008 (Stockman et al., 2008). One possible reason for this change is that in January of 2023, the Council on Academic Accreditation for Audiology and Speech-Language Pathology (CAA), the organization that accredits American SLP programs, revised their accreditation standards to include a separate requirement that institutions must provide evidence that diversity, equity and inclusion are embedded into academic and clinical coursework, in theory and practice (CAA, n.d.). This new standard also includes specific reference to development of cultural and linguistic competence (CAA, n.d.). Additionally, ASHA’s Office of Multicultural Affairs hosts a faculty resource page (e.g., sample syllabi and instructional activities) for curricular infusion and dedicated CLD courses (ASHA, n.d.). Given the many cultural and linguistic similarities between diverse countries like Canada, Australia and the United Kingdom and the United States, other institutions and faculty may wish to consult these ASHA resources, or others such as Becoming Culturally Responsive, (a site that provides pedagogical materials on CLD practice) when looking for ways to increase the CLD competence of their future students (Becoming Culturally Responsive, n.d.). Such efforts may result in students feeling better prepared to provide service delivery in languages other than English or French.

Unfortunately, even with combined curricular infusion and dedicated CLD coursework, an additional challenge will likely remain; the paucity of research conducted in languages other than English, French and Spanish. Little is known about speech and language development milestones and manifestation of disorders for monolingual and bilingual speakers of other languages. There are minimal assessment tools for speakers of heritage languages, and little research into effective intervention practices or development of intervention materials for speakers of other commonly spoken languages in Canada. For instance, in Canada, the number of people who speak Punjabi rose 49% between 2016 and 2021, and there are now more than a half million individuals who speak Punjabi at home (Statistics Canada, 2022i). However, Canadian SLP course instructors seeking to integrate information about Punjabi speech and language development, assessment or intervention would be confronted with a dearth of information when looking for relevant studies to integrate into coursework. Fulsome integration of contextually appropriate CLD service delivery issues in SLP programs is therefore dependent on high-quality research into languages other than English, French and Spanish.

### Sex and gender

Canadian SLP students are overwhelmingly cisgender females (96%). These findings are consistent with those documented in previous demographic surveys of SLPs and SLP students in Canada and abroad (ASHA, 2023; CASLPO, 2024; RCSLT, 2023; SAC, 2021). In terms of sex, this is evidently not representative of the Canadian population, where 50.9% are females (Statistics Canada, 2022a). Regarding gender, 99.77% of Canadian identified as cisgender, and only 0.33% as trans or nonbinary in the population at large across all age groups (Statistics Canada, 2022c). However, the proportion of trans or nonbinary individuals aged 20-24 in Canada was higher, at 0.85% (Statistics Canada, 2022c). Given that the mean age of our sample was 25, and 1.3% identified as trans or nonbinary, the trans and nonbinary community is well represented in the current Canadian SLP student population.

The implications for clinicians and clients of this sex and gender imbalance in the profession have been discussed in prior literature. Speech-Language Pathology is described as a *caring or helping* profession, that pertains to *communication*, skills that have historically been considered “female,” leading to the perception that the profession is “women’s work” (Litosseliti & Leadbeater, 2013). Costello (2005) argues that prospective male students may be discouraged from considering ‘female’ professions, such as speech-language pathology, because integrating one’s professional identity into their gender identity is easier when the two are consistent. Indeed, male SLPs report obstacles in navigating a “female” profession (Azios & Bellon-Harn, 2021). These include challenges with other’s perception of their performance of stereotypically female roles (e.g., playing and talking with children); feeling alone or isolated (e.g., being the only male in their academic cohort or their workplace); and dealing with others’ uncertainty as to whether they are suitable for the career (e.g., questions as to why they did not consider careers in more stereotypically “male” rehabilitation professions such as occupational or physical therapy). Skeat et al. (2022) argue that changing the sex and gender composition of the profession will require more than simply encouraging males to apply. They posit that this type of change will require a more in-depth consideration of gendered discourses in the profession and their implications on professional practice (Skeat et al., 2022). It is possible that as gender roles continue to be contested and change in society at large, there may be greater representation of men in helping professions, such as Speech-Language Pathology.

Gender can also determine the SLP’s perception of their clients. For instance, boys have long been identified with communication and academic difficulties and placed in special education programs at higher rates than girls (e.g., Dever et al., 2016; Wehmeyer & Schwarz, 2001). It has been suggested that gender bias, (e.g., stereotypical favourable *or* prejudiced beliefs about an individual based on their gender) may play a role in this phenomenon (Wehmeyer & Schwarz, 2001). To our knowledge, limited research on this topic has been conducted in the field of speech-language pathology. However, studies of teacher perceptions have found that girl students are generally perceived more favourably in terms of temperament and educational competence than boys by both men and women teachers (e.g.,Mullola et al., 2012). However, men teachers perceived this gap between genders to be smaller than women teachers, and perceived boy students more positively than their women teachers did (Mullola et al., 2012). These gendered perceptions can have negative impacts for both boy and girl students, who may experience over and under identification of communication and academic-related challenges respectively. Teaching is also a stereotypically “female” profession with considerable overlap in pediatric speech-language pathology (e.g., evaluating and providing instruction in academic and language-related skills of children) and it is not implausible that these same gender biases exist in the field of SLP.

### Socio-economic status (SES)

Most Canadian SLP students come from high socio-economic status households. In Canada at large, the median household income in 2021 was $68,400.00 (Statistics Canada, 2023). Conversely, nearly three quarters of the students who answered the survey question about estimated total household income reported that their household income was greater than $75,000.00 (72%, N=318). It is unlikely that SLP students are earning this type of salary independently while completing an intensive Master’s program. The students surveyed also reported that this total household income supported on average 3.5 individuals. These findings suggest that most SLP students are supported financially by their caregivers while studying and that most SLP students’ caregivers are earning total incomes higher than the Canadian median.

A recent survey of Canadian graduate students across research and course-based disciplines found that MSc and PhD students experience financial stress, are burdened by tuition payments, and often must take on extra work when studying to support themselves (Laframboise et al., 2023). Clinical SLP programs are no different. Applications are costly (Kovacs, 2022) and tuition is expensive, costing upwards of $25,000.00 total (e.g., University of Toronto, n.d.b). Clinical SLP programs are also fast-paced, and students typically have 4-5 courses concurrently, along with assignments, readings and clinical placements. This can make it challenging to maintain part-time employment while studying.^2^ Evidence also suggests that these financial barriers affect graduate students from underrepresented communities more deeply (Laframboise et al., 2023).

### Racial, ethnic and cultural diversity

Canadian SLP students reported 1084 total and 151 unique racial and ethnic identities, most of which were location-based. North American and European racial/ethnic origins were overwhelmingly the most reported (40.3% and 22.1% respectively). Not all participants elected to identify themselves by physical characteristics like skin colour, but of those who did most reported being Caucasian (11.9%). Following North American and European origins, the next largest racial/ethnic group was Asian at 10.5% of respondents. Of those, most were South Asian (4% of the total), or Chinese (2% of the total). Only 1.7% of respondents identified as Indigenous, and 0.8% of respondents identified as black. These results are in contrast with the most recent data collected by Statistics Canada (Statistics Canada, 2022e). In the 2021 census, more than 450 unique origins were reported by Canadians, and though close to 70% of Canada’s population was white, 7.1% were South Asian, 4.7% were Chinese, 5% were Indigenous and 4.3% were black (Statistics Canada, 2022e; h). These findings suggest that Canada’s SLP students are primarily of North American and European racial and ethnic origins and are not wholly representative of all racial groups in Canada. The differences between SLP students and the Canadian population are greatest for black and Indigenous communities.

Culturally, results were similar, with 121 unique cultural identities reported, and the largest proportion of respondents indicating they were culturally Canadian (47.4%). This is consistent with Statistics Canada data, where “Canadian” was also the most selected cultural origin, but the proportion of individuals who identified this way in Canada was much smaller than in the SLP student population, at 15.6%. (Statistics Canada, 2022e) Some SLP student respondents elected to report their religion as a racial/ethnic or cultural identity. The most reported religions were Christian, Catholic, Jewish, and Islam (Muslim origins). This is largely consistent with Statistics Canada data, where common religious affiliations were Christian (53.3%), Muslim (4.9%) and Jewish (1.1%) (Statistics Canada, 2022e). Though Sikhism and Hinduism were on the rise in Canada, now comprising 2.1% and 2.3% of the Canadian population, these religions were reported by only 1 and 2 respondents respectively in our data (Statistics Canada, 2022e). Notable cultural groups not recognized in racial/ethnic identity results of this survey included “Desi” and “Latin American/Latin/Latinx”. Desi was not an identity reported by Statistics Canada, though 2.3% of Canadian SLP students identified with this term. The proportion of Latin SLP students (1%) is similar to that of the Canadian population (1.6%). Culturally, only 0.8% of respondents identified as Indigenous though there are nearly 2.2 million Canadians (5%) who reported Indigenous ancestry in the 2021 census (Statistics Canada, 2022h).

A recent survey of Canadian medical students identified a similar trend in the representation of both black and Indigenous students. Regarding the black population, it was found that 1.7% of Canadian medical students identified as black, compared to 4.3% of Canadian census respondents (Khan et al., 2020; Statistics Canada, 2022e). These medical students face intrinsic and extrinsic barriers through self-doubt, financial barriers, systemic racism and biased institutional policies, as well as challenges with belonging (Egbedeyi et al., 2022). For example, an accountability report compiled by the University of Toronto’s Temerty Faculty of Medicine found that 56% of Black medical students, 66% of Black residents and 39% of Black fellows experienced discrimination at least once within that past year (University of Toronto, 2022). Black students face barriers to higher education at every part of the process from success in undergraduate programs to gaining relevant experience and mentorship and application to graduate programs to finding role models in healthcare that understand their unique experiences.

Additionally, Khan et al. (2020) found that 3.5% of Canadian medical students identified as Indigenous, compared to Canada’s reported Indigenous population of 5% (Statistics Canada, 2022h). Of this 3.5%, it appears as though there is a further disparity between those that live in urban settings and those who live on reserves or in rural or Northern communities (Henderson et al., 2021). Those who do not live in urban areas often face challenges such as under-funded and ill-equipped high school education, which impacts their ability to successfully apply to undergraduate programs, let alone graduate programs such as SLP or medical school. Specialized application streams, self-identification, and diversity quotas only serve to homogenize the Indigenous community and bias acceptances towards those that most resemble what may be considered the ‘norm’ of applicants regarding other factors such as geographical location and socioeconomic status (Brisebois & Cardinal, 2024). Indigenous students face unique challenges in their pursuit of higher education, challenges that are still present once within graduate programs. Similarly, to black students, Indigenous students face systemic racism, financial barriers, feelings of self-doubt and shame, discrimination from faculty, peers, and clients, as well as lack of representation. Canadian universities have been slow to adopt the collection of race-based data from their students, but this data is a key component of understanding racial disparities and their impacts in educational settings.

## Conclusions

Findings from this nationwide survey of clinical SLP students suggest that the profession is trending toward greater diversity in terms of bi and multilingualism, trans and nonbinary representation and representation of some but not all racial/ethnic and cultural groups. Results highlight the need for greater representation of males, black and Indigenous students, those from lower SES backgrounds and students whose native languages are not English or French. All prospective students, regardless of their unique intersecting identities, should have equal opportunity to access SLP graduate programs, not only because access to education is a right that should not be restricted by language, race or other demographic factors, but also because greater diversity in our clinical cohorts will lead to enriched learning experiences for SLP students and better outcomes for our clients. Diversity in higher education classrooms can lead to better educational outcomes (Gottfredson et al., 2008), higher GPAs (Lau, 2022) and improved critical thinking and problem-solving skills (Hurtado, 2001). Diverse classroom and campus experiences can also result in increased cultural awareness and provide students opportunities to learn how to work with individuals who are different from them (Meacham et al., 2003).

In terms of the clinical implications of increased diversity, patients report that concordance, where patient and practitioner share a common language, race or culture, is an effective method for improving cultural competence and patient outcomes in healthcare (e.g., Cuevas et al., 2017; Handtke et al., 2019). A recent systematic review found that linguistic concordance in healthcare resulted in increased access to care and information, patient satisfaction, improved interpersonal relationships and fewer missed appointments (Lor & Martinez, 2020). There is also a growing body of research suggesting that racial concordance can result in improved healthcare and therapeutic relationships (e.g., Jetty et al., 2022). Though there is little research in speech-language pathology examining the effect of concordance, it is likely that these intersecting factors would have an impact, perhaps to an even greater extent, given the focus on assessment and intervention in language and communication, skills mediated by race and culture. Future research should explore this possibility. Finally, increased clinical diversity would also result in increased diversity knowledge in the field and reduce burdens for minority SLPs. For instance, at present, only 3 of the 525 surveyed SLPs students felt competent to provide clinical service delivery in Mandarin, despite it being the third most spoken language in Canada. When an English or French speaking SLP encounters a Mandarin client in practice, they may, if they are lucky, have one Mandarin speaking colleague to turn to. If not, they must rely on scant research literature available on bilingualism in heritage languages, or on other cultural linguistic informants like family or community members. Increasing the number of Mandarin speaking SLPs means greater access to linguistic and cultural knowledge for all SLPs, and reduced burden for minority SLPs who may have previously been the lone holder of this knowledge in their workplace. In summary, increasing the diversity of the clinical SLP student population should be prioritized to ensure equitable access to higher education and greater outcomes for SLP clients.

### Future Directions and Recommendations for Diversification

Given the immeasurable and increasing diversity in countries like Canada, the United States, Australia and the United Kingdom, it is unlikely that we will ever achieve 1:1 representation of all intersecting identities in our respective clinical populations. However, there are steps that can be taken take to promote diversity in clinical education programs, and consequently the profession, both in Canada and elsewhere. First, reducing financial barriers to entry. At present, in the Canadian context, low SES students are underrepresented in SLP programs. SES is related to newcomer status, and accordingly to linguistic, racial and cultural identity (Statistics Canada, 2022f). Increased funding opportunities, scholarships or bursaries for professional and course-based master’s programs are one potential solution. Other options for minimizing financial barriers include waiving partial fees for students from low SES backgrounds or offering part-time clinical education programs to allow students to work and support themselves. Second, SLP programs in Canada and abroad, could consider removing language proficiency requirements like completion of the TOEFL for non-native English or French speaking students to reduce barriers for these students and increase linguistic diversity, an approach has already been employed at some Canadian institutions. Third, to better prepare all students for engaging in clinical practice with culturally and linguistically diverse students, institutions could make use of existing resources that address how to infuse cultural and linguistic competence into domain-specific coursework, and on how to establish CLD specific courses that teach domain-general skills (e.g., cultural humility, dynamic assessments or working with interpreters).

We also advocate for all clinical education programs and for governing bodies to collect and report demographic data about SLP students and practicing SLPs on an ongoing basis. This will allow us to gauge whether any efforts to diversify profession are having long-term impacts, and where efforts need to be focused on in the future. Finally, we acknowledge increase the diversity of SLP students and SLPs will not be sufficient on its own. It is also critical that researchers engage with languages other than English, French and Spanish, because without high quality research into speech and language development, assessment, and intervention in other heritage languages, instructors cannot embed this knowledge into their courses and clinicians cannot use this information in practice.

## Supporting information

Supplemental File 2 (Survey)

## Data Availability

All data produced in the present study are available upon reasonable request to the authors

## Acknowledgements

This project is supported by the Canada Graduate Scholarship – Doctoral Program (CGS-D; Social Sciences and Humanities Research Council of Canada) awarded to EW, and the Social Sciences and Humanities Research Council of Canada (SSHRC) Insight Grant (#435-2024-0713) awarded to MM.

## Footnotes

1 Notable exceptions are Dalhousie and Laurentian University. Dalhousie’s admission requirements page explicitly states because the school values diversity, students whose native language is not English will not have to prove their proficiency through a formal standardized test . However, both Laurentian and Dalhousie, and all other programs, also state that high or native levels of English or French proficiency are required to successfully complete their programs, which may discourage non-native English or French students from applying

2 Students may apply for loans or scholarships and bursaries to support themselves financially during graduate school. However, there are limited scholarship opportunities that provide substantial funding to clinical graduate students. This means the competition for these awards is fierce, and many students are left only with the option to take out additional student loans if their families are not able to support them.

