## Supplemental Table 1 for "Who are the Speech-Language Pathologists of the Future? Results of a national demographic survey of Canadian SLP Students"

**Appendix Table 1.**

*Overall response rate by institution and year of study*

|  | Institution | N Respondents /N Enrolled Students | Response Rate |
| --- | --- | --- | --- |
|  | Dalhousie Y1 | 21/24 | 88% |
|  | Dalhousie Y2 | 19/23 | 83% |
|  | Dalhousie Y3 | 4/22 | 18% |
|  | Dalhousie Total | 44/69 | 64% |
|  | Laurentian Y1 | 2/6 | 33% |
|  | Laurentian Y2 | 4/9 | 44% |
|  | Laurentian Total | 6/15 | 40% |
|  | McGill Y1 | 23/30 | 77% |
|  | McGill Y2 | 29/30 | 96% |
|  | McGill Total | 52/60 | 86% |
|  | McMaster Y1 | 11/36 | 31% |
|  | McMaster Y2 | 21/32 | 66% |
| McMaster Total | | 32/68 | 47% |
|  | University of Alberta Y1 | 35/70 | 50% |
|  | University of Alberta Y2 | 21/60 | 35% |
|  | University of Alberta Total | 59/135 | 44% |
|  | University of British Columbia Y1 | 16/38 | 42% |
|  | University of British Columbia Y2 | 22/36 | 61% |
|  | University of British Columbia Total | 38/74 | 51% |
|  | Université de Laval Y1 | 13/50 | 26% |
|  | Université de Laval Y2 | 16/50 | 32% |
|  | Université de Laval total | 29/100 | 29% |
|  | Université de Montréal Y1 | 37/72 | 51% |
|  | Université de Montréal Y2 | 28/71 | 39% |
|  | Université de Montréal Total | 65/144 | 44% |
|  | University of Ottawa Y1 | 12/25 | 48% |
|  | University of Ottawa Y2 | 5/25 | 25% |
|  | University of Ottawa Total | 17/50 | 32% |
|  | Université de Québec à Trois Rivières Y1 | 20/24 | 83% |
|  | Université de Québec à Trois Rivières Y2 | 18/22 | 82% |
|  | Université de Québec à Trois Rivières Total | 38/45 | 84% |
|  | University of Toronto Y1 | 33/57 | 57% |
|  | University of Toronto Y2 | 38/68 | 56% |
|  | University of Toronto Total | 71/125 | 57% |
|  | Western University Y1 | 39/54 | 72% |
|  | Western University Y2 | 31/50 | 62% |
|  | Western University Total | 76/104 | 72% |
|  | University Total | **525/987** | **53%** |
