## Supplemental File 2 (Survey) for "Who are the Speech-Language Pathologists of the Future? Results of a national demographic survey of Canadian SLP Students"

### Student Demographic Survey

Please complete the survey below.

Thank you!

| | |
|---|---|
| To proceed in English, select English from the menu.<br><br>Pour continuer en français, sélectionnez français dans le menu. | ◯ English<br>◯ Français |
| What institution are you currently enrolled in? | ◯ Dalhousie University<br>◯ Laurentian University<br>◯ Laval University<br>◯ McGill University<br>◯ McMaster University<br>◯ Quebec a Trois Rivieres<br>◯ University of Alberta<br>◯ University of British Columbia<br>◯ University of Montreal<br>◯ University of Ottawa<br>◯ University of Toronto<br>◯ Western University<br>◯ Prefer not to disclose |
| When are you scheduled to graduate? | ◯ 2024<br>◯ 2025<br>◯ 2026<br>◯ Prefer not to disclose |
| What is your current age? If you prefer not to disclose, please type N/A. | __________________________ |
| What is your sex? (sex here is defined as the sex assigned at birth, typically based on a person's reproductive system and physical characteristics). | ◯ Male<br>◯ Female<br>◯ Prefer not to disclose |
| What is your gender identity? (gender here is defined as the gender that a person internally feels and/or the gender that a person publicly expresses in their daily life e.g., nonbinary, cisgender woman). If you prefer not to disclose, please type N/A. | __________________________ |
| How many languages do you speak? | ◯ 1<br>◯ 2<br>◯ 3+ |
| What is your first language? | __________________________ |
| In what environment did you learn your first language? You may select more than one option. | ☐ Home<br>☐ School<br>☐ Formal instruction (e.g., a language class or after-school program)<br>☐ In a country where the language was spoken<br>☐ Other<br>☐ Prefer not to disclose |

| | |
|---|---|
| You selected other, please specify: | __________________________________ |
| Are you sufficiently proficient in this language to be able to provide speech-language pathology services upon graduation? | ◯ Yes<br>◯ No |
| What is your second language? | __________________________________ |
| In what environment did you learn your second language? You may select more than one option. | ☐ Home<br>☐ School<br>☐ Formal instruction (e.g., a language class or after-school program)<br>☐ In a country where the language was spoken<br>☐ Other<br>☐ Prefer not to disclose |
| You selected other, please specify: | __________________________________ |
| Are you sufficiently proficient in this language to be able to provide speech-language pathology services upon graduation? | ◯ Yes<br>◯ No |
| What is your third language? | __________________________________ |
| In what environment did you learn your third language? You may select more than one option. | ☐ Home<br>☐ School<br>☐ Formal instruction (e.g., a language class or after-school program)<br>☐ In a country where the language was spoken<br>☐ Other<br>☐ Prefer not to disclose |
| You selected other, please specify: | __________________________________ |
| Are you sufficiently proficient in this language to be able to provide speech-language pathology services upon graduation? | ◯ Yes<br>◯ No |
| What language(s) are most used to communicate in your home environment? | __________________________________ |

Please indicate the racial ethnic background(s) with which you identify. You may select more than one response.

- ☐ Abenaki
- ☐ Acadian
- ☐ Afar
- ☐ Afghan
- ☐ African
- ☐ African American
- ☐ African Canadian
- ☐ African Caribbean
- ☐ African Nova Scotian
- ☐ Afrikaner
- ☐ Ahousaht
- ☐ Akan
- ☐ Albanian
- ☐ Albertan
- ☐ Algerian
- ☐ Algonquin
- ☐ Alsatian
- ☐ American
- ☐ Amhara
- ☐ Amish
- ☐ Anglo-Indian
- ☐ Angolan
- ☐ Anguillan
- ☐ Anishinaabe
- ☐ Antiguan
- ☐ Apache
- ☐ Arab
- ☐ Arawak
- ☐ Argentinian
- ☐ Armenian
- ☐ Aruban
- ☐ Ashanti
- ☐ Asian
- ☐ Assiniboine
- ☐ Assyrian
- ☐ Atikamekw
- ☐ Australian
- ☐ Austrian
- ☐ Azerbaijani
- ☐ Azorean
- ☐ Bahamian
- ☐ Bahraini
- ☐ Baloch
- ☐ Bambara
- ☐ Bamileke
- ☐ Bangladeshi
- ☐ Bantu
- ☐ Baoulé
- ☐ Barbadian
- ☐ Bashkir
- ☐ Basque
- ☐ Batswana
- ☐ Bavarian
- ☐ Beaver (Dunne-za)
- ☐ Belgian
- ☐ Belizean
- ☐ Bengali
- ☐ Beninese
- ☐ Berber
- ☐ Bermudian
- ☐ Bhutanese
- ☐ Black
- ☐ Blackfoot
- ☐ Blood (Kainai)
- ☐ Bohemian
- ☐ Bolivian
- ☐ Bosniak
- ☐ Bosnian
- ☐ Brazilian

- ☐ Breton
- ☐ British
- ☐ British Columbian
- ☐ Bruneian
- ☐ Buddhist
- ☐ Bulgarian
- ☐ Burkinabe
- ☐ Burmese
- ☐ Burundian
- ☐ Byelorussian
- ☐ Cambodian
- ☐ Cameroonian
- ☐ Canadian
- ☐ Cape Bretoner
- ☐ Cape Verdean
- ☐ Carib
- ☐ Caribbean
- ☐ Carrier (Dakelh)
- ☐ Catalan
- ☐ Caucasian (White)
- ☐ Caymanian
- ☐ Cayuga
- ☐ Celtic
- ☐ Central African
- ☐ Central African
- ☐ Central American
- ☐ Central American Indian (Indigenous)
- ☐ Central Asian
- ☐ Chadian
- ☐ Chaldean
- ☐ Channel Islander
- ☐ Chechen
- ☐ Chemainus (Stz'uminus)
- ☐ Cherokee
- ☐ Cheyenne
- ☐ Chilcotin (Tsilhqot'in)
- ☐ Chilean
- ☐ Chin
- ☐ Chinese
- ☐ Chipewyan (Denesuline)
- ☐ Choctaw
- ☐ Christian
- ☐ Circassian
- ☐ Coast Salish
- ☐ Colombian
- ☐ Comorian
- ☐ Congolese
- ☐ Coptic
- ☐ Cornish
- ☐ Corsican
- ☐ Costa Rican
- ☐ Cowichan
- ☐ Cree
- ☐ Creole
- ☐ Croatian
- ☐ Crow
- ☐ Cuban
- ☐ Cypriot
- ☐ Czech
- ☐ Czechoslovakian
- ☐ Dakota
- ☐ Danish
- ☐ Delaware (Lenape)
- ☐ Dene
- ☐ Dene Tha' (Slavey)
- ☐ Dinka
- ☐ Ditidaht
- ☐ Djiboutian
- ☐ Dominica Islander
- ☐ Dominican
- ☐ Doukhobor

- ☐ Dutch
- ☐ Dzawada'enuxw
- ☐ East African
- ☐ East Asian
- ☐ Eastern European
- ☐ Ecuadorian
- ☐ Edo
- ☐ Egyptian
- ☐ Ehattesaht
- ☐ English
- ☐ Eritrean
- ☐ Esan
- ☐ Estonian
- ☐ Ethiopian
- ☐ Eurasian
- ☐ European
- ☐ Ewe
- ☐ Fante
- ☐ Faroese
- ☐ Fijian
- ☐ Filipino
- ☐ Finnish
- ☐ First Nations
- ☐ Flemish
- ☐ Franco Ontarian
- ☐ French
- ☐ French Canadian
- ☐ Frisian
- ☐ Fulani
- ☐ Ga-Adangbe
- ☐ Gabonese
- ☐ Galician
- ☐ Gambian
- ☐ Gaspesian
- ☐ Georgian
- ☐ German
- ☐ Ghanaian
- ☐ Gibraltarian
- ☐ Gitxsan
- ☐ Goan
- ☐ Greek
- ☐ Greek Cypriot
- ☐ Greenlandic
- ☐ Grenadian
- ☐ Guadeloupean
- ☐ Guatemalan
- ☐ Guinean
- ☐ Gujarati
- ☐ Guyanese
- ☐ Gwa'sala
- ☐ Gwich'in
- ☐ Haida
- ☐ Haisla
- ☐ Haitian
- ☐ Halalt
- ☐ Hän (Tr'ondëk Hwëch'in)
- ☐ Harari
- ☐ Hausa
- ☐ Hawaiian
- ☐ Hazara
- ☐ Heiltsuk
- ☐ Hesquiaht
- ☐ Hindu
- ☐ Hispanic
- ☐ Hmong
- ☐ Homalco
- ☐ Honduran
- ☐ Hong Konger
- ☐ Huguenot
- ☐ Hungarian
- ☐ Huron (Wendat)

- ☐ Hutterite
- ☐ Hutu
- ☐ Huu-ay-aht
- ☐ Icelandic
- ☐ Igbo
- ☐ Igorot
- ☐ Ilocano
- ☐ Indian (India)
- ☐ Indo-Caribbean
- ☐ Indo-Fijian
- ☐ Indo-Guyanese
- ☐ Indonesian
- ☐ Innu
- ☐ Interior Salish
- ☐ Inuit
- ☐ Inuvialuit
- ☐ Iranian
- ☐ Iraqi
- ☐ Irish
- ☐ Iroquois (Haudenosaunee)
- ☐ Israeli
- ☐ Italian
- ☐ Ivorian
- ☐ Jamaican
- ☐ Japanese
- ☐ Jatt
- ☐ Javanese
- ☐ Jewish
- ☐ Jordanian
- ☐ K'omoks
- ☐ Kabyle
- ☐ Karen
- ☐ Kashmiri
- ☐ Kashubian
- ☐ Kaska
- ☐ Kazakh
- ☐ Kenyan
- ☐ Khmer
- ☐ Kikuyu
- ☐ Kittitian/Nevisian
- ☐ Korean
- ☐ Kosovar
- ☐ Ktunaxa (Kutenai)
- ☐ Kurdish
- ☐ Kuwaiti
- ☐ Kwakiutl
- ☐ Kwakwa̱ka̱’wakw
- ☐ Kyrgyz
- ☐ Kyuquot/Cheklesaht
- ☐ Laich-kwil-tach
- ☐ Lakota
- ☐ Laotian
- ☐ Latin American
- ☐ Latvian
- ☐ Lebanese
- ☐ Lekwungen
- ☐ Liberian
- ☐ Libyan
- ☐ Liechtensteiner
- ☐ Lithuanian
- ☐ Luba
- ☐ Luo
- ☐ Luxembourger
- ☐ Macedonian
- ☐ Maghrebi
- ☐ Maharashtrian
- ☐ Malagasy
- ☐ Malahat
- ☐ Malawian
- ☐ Malay
- ☐ Malayali

- ☐ Malaysian
- ☐ Malian
- ☐ Malinké
- ☐ Maliseet
- ☐ Maltese
- ☐ Mamalilikulla
- ☐ Manitoban
- ☐ Manx
- ☐ Maori
- ☐ Mapuche
- ☐ Maroon
- ☐ Martinican
- ☐ Mauritanian
- ☐ Mauritian
- ☐ Mayan
- ☐ Mennonite
- ☐ Métis
- ☐ Mexican
- ☐ Mi'kmaq
- ☐ Middle Eastern
- ☐ Mohawk
- ☐ Moldovan
- ☐ Mongolian
- ☐ Montagnais
- ☐ Montenegrin
- ☐ Montserratan
- ☐ Moose Cree
- ☐ Moravian
- ☐ Moroccan
- ☐ Mossi
- ☐ Mowachaht/Muchalaht
- ☐ Mozambican
- ☐ Muslim
- ☐ Musqueam
- ☐ Nakwaxda'xw
- ☐ Namgis
- ☐ Namibian
- ☐ Naskapi
- ☐ Navajo
- ☐ Ndebele
- ☐ Nepali
- ☐ New Brunswicker
- ☐ New Zealander
- ☐ Newfoundlander
- ☐ Nez Perce
- ☐ Nicaraguan
- ☐ Nigerian
- ☐ Nigerien
- ☐ Nisga'a
- ☐ Nlaka'pamux (Thompson)
- ☐ Norman
- ☐ North African
- ☐ North American
- ☐ Northern European
- ☐ Northern Irish
- ☐ Norwegian
- ☐ Nova Scotian
- ☐ Nubian
- ☐ Nuchatlaht
- ☐ Nuu-chah-nulth
- ☐ Nuxalk
- ☐ Oceanian
- ☐ Odawa
- ☐ Oji-Cree
- ☐ Ojibway
- ☐ Okanagan (Syilx)
- ☐ Omani
- ☐ Oneida
- ☐ Onondaga
- ☐ Ontarian
- ☐ Orcadian

- ☐ Oromo
- ☐ Pacific Islander
- ☐ Pakistani
- ☐ Palestinian
- ☐ Panamanian
- ☐ Papua New Guinean
- ☐ Paraguayan
- ☐ Pashtun
- ☐ Passamaquoddy
- ☐ Penelakut
- ☐ Pennsylvania Dutch
- ☐ Persian
- ☐ Peruvian
- ☐ Piikani
- ☐ Pipil
- ☐ Plains Cree
- ☐ Polish
- ☐ Polynesian
- ☐ Portuguese
- ☐ Potawatomi
- ☐ Prince Edward Islander
- ☐ Puerto Rican
- ☐ Punjabi
- ☐ Qalipu Mi'kmaq
- ☐ Quatsino
- ☐ Québécois
- ☐ Quechua
- ☐ Réunionnais
- ☐ Rohingya
- ☐ Roma
- ☐ Romanian
- ☐ Russian
- ☐ Ruthenian
- ☐ Rwandan
- ☐ Sahtú (North Slavey)
- ☐ Saint Helenian
- ☐ Salish
- ☐ Salvadorean
- ☐ Sami
- ☐ Samoan
- ☐ Saskatchewanian
- ☐ Saudi Arabian
- ☐ Saulteaux
- ☐ Scandinavian
- ☐ Scottish
- ☐ Secwepemc (Shuswap)
- ☐ Seneca
- ☐ Senegalese
- ☐ Serbian
- ☐ Serer
- ☐ Seychellois
- ☐ Shawnee
- ☐ Shishalh (Sechelt)
- ☐ Shona
- ☐ Sicilian
- ☐ Sierra Leonean
- ☐ Sikh
- ☐ Siksika
- ☐ Sindhi
- ☐ Singaporean
- ☐ Sinhalese
- ☐ Sioux
- ☐ Slavic
- ☐ Slovak
- ☐ Slovenian
- ☐ Snuneymuxw
- ☐ Somali
- ☐ Soninke
- ☐ South African
- ☐ South American
- ☐ South American Indian (Indigenous)

- ☐ South Asian
- ☐ South Sudanese
- ☐ Southeast Asian
- ☐ Southeast European
- ☐ Southern European
- ☐ Spanish
- ☐ Squamish
- ☐ Sri Lankan
- ☐ St. Lucian
- ☐ St'at'imc
- ☐ (Lillooet) Stó:lō
- ☐ Stoney (Nakoda)
- ☐ Sudanese
- ☐ Surinamese
- ☐ Swahili
- ☐ Swampy Cree
- ☐ Swazi
- ☐ Swedish
- ☐ Swiss
- ☐ Syrian
- ☐ T'Sou-ke
- ☐ Tagish
- ☐ Tahitian
- ☐ Tahltan
- ☐ Taiwanese
- ☐ Tajik
- ☐ Tamil
- ☐ Tanzanian
- ☐ Tatar
- ☐ Telugu
- ☐ Thai
- ☐ Tibetan
- ☐ Tigrian
- ☐ Tla-o-qui-aht
- ☐ Tla'amin (Sliammon)
- ☐ Tlatlasikwala
- ☐ Tlicho (Dogrib)
- ☐ Tlingit
- ☐ Tlowitsis
- ☐ Togolese
- ☐ Tongan
- ☐ Transylvanian
- ☐ Trinidadian/Tobagonian
- ☐ Tsek'ene (Sekani)
- ☐ Tseshaht
- ☐ Tsimshian
- ☐ Tsuu T'ina (Sarcee)
- ☐ Tswana
- ☐ Tunisian
- ☐ Turkish
- ☐ Turkish Cypriot
- ☐ Turkmen
- ☐ Tuscarora
- ☐ Tutchone
- ☐ Tutsi
- ☐ Ucluelet
- ☐ Ugandan
- ☐ Ukrainian
- ☐ Ulster Scot
- ☐ United Empire Loyalist
- ☐ Uruguayan
- ☐ Uyghur
- ☐ Uzbek
- ☐ Venezuelan
- ☐ Vietnamese
- ☐ Vincentian
- ☐ Walloon
- ☐ Welsh
- ☐ West African
- ☐ West Asian
- ☐ West Indian

| | |
|---|---|
| | ☐ Western European<br>☐ Wet'suwet'en<br>☐ Wolof<br>☐ Woodland Cree<br>☐ W̱SÁNEĆ (Saanich)<br>☐ Wuikinuxv<br>☐ Xhosa<br>☐ Yazidi<br>☐ Yemeni<br>☐ Yoruba<br>☐ Yugoslavian<br>☐ Zambian<br>☐ Zimbabwean<br>☐ Zoroastrian<br>☐ Zulu<br>☐ Prefer not to disclose |
| Is there anything more you'd like to share about your race or ethnicity? (For example, one researcher would state that she is Gujarati, another would state she is Caucasian). | ________________________________ |
| What cultures do you identify with? (i.e. Canadian, Jewish-Orthodox, Desi-Canadian, etc.)<br><br>If you would like to see more examples visit this link: https://www12.statcan.gc.ca/census-recensement/2021/ref/questionnaire/ancestry.cfm<br><br>If you prefer not to disclose, please type N/A. | ________________________________ |
| Do you consider yourself, first, second, or third + generation Canadian? (Here, first generation Canadian is defined as an individual whose parents or caregivers were not born in Canada, second generation is an individual for whom at least one parent or caregiver was born in Canada, and third generation is defined as an individual for whom at least one grandparent was born in Canada). | ◯ First generation Canadian<br>◯ Second generation Canadian<br>◯ Third+ generation Canadian<br>◯ Prefer not to disclose |
| Please estimate your family's total household income for 2023. This includes your parents/caregivers income, your income, or your siblings. | ◯ 0 - 5,000.00<br>◯ 5,000.00 - 10,000.00<br>◯ 10,000.00- 25,000.00<br>◯ 25, 000.00 - 50,000.00<br>◯ 50, 000.00 - 75, 000.00<br>◯ 75, 000.00 - 100,000.00<br>◯ 100,000.00 - 150, 000.00<br>◯ 150,000.00 - 200, 000.00<br>◯ 200,000.00 - 250, 000.00<br>◯ 250,000.00 +<br>◯ Prefer not to disclose |
| Please indicate how many people this total household income supports (e.g., a family with two caregivers and two children would equate to 4 individuals supported by the total household income). If you prefer not to disclose, please type N/A. | __________________________ |
| Please provide the first three characters of your postal code (e.g., if your postal code in M5G 1V7, you would input "M5G"). If you prefer not to disclose, please type N/A. | __________________________ |
| Are there any questions or concerns you have regarding today's survey? For example, were there any questions above where you felt unsatisfied with the options you | ________________________________ |
